# Plasma Biomarkers of Acute Respiratory Distress Syndrome Vary with Body Size in the Absence of Acute Illness

**DOI:** 10.64898/2026.09.19.26362333

**Authors:** Timothy G. Gaulton, Lorenzo Berra, Maurizio Cereda

**Author notes:** Corresponding author. Timothy G. Gaulton, Mass General Brigham, Department of Anesthesiology, 275 Charles Street, Warren Building 1225D, Boston, MA 02114, United States.

## Abstract

**Rationale:** Plasma proteins are used as biomarkers of inflammation and injury in acute respiratory distress syndrome (ARDS). Whether they vary with body size before ARDS is not well established.

**Objectives:** To determine whether ARDS biomarkers vary with body mass index in adults without acute illness.

**Methods:** We analyzed 8,680 ambulatory adults in a cross sectional study of the All of Us Research Program with proteomics covering 5,420 protein assays. We grouped 35 proteins associated with ARDS risk or mortality into five compartments by the pathophysiological role assigned in a published systematic review, registered in advance. We modeled body mass index as a restricted cubic spline and reported the difference in each protein between 35 and 25 kg/m^2^ in standard deviations, adjusted for age, sex, smoking, thirteen chronic conditions and ten genetic principal components. Each estimate was ranked against all 5,420 assays.

**Results:** Median age was 50 years and median body mass index 27.2 kg/m2. Associations with body mass index across all assays were predominantly positive, median 0.065 standard deviations. All four epithelial proteins were lower at higher body mass index, each below at least 92% of assays. Soluble receptor for advanced glycation end products was lower by 0.350 standard deviations and surfactant protein A2, surfactant protein D and club cell secretory protein 16 were lower by 0.081 to 0.084 standard deviations. Interleukin-6 and E-selectin were 0.534 and 0.465 standard deviations higher, above 99% of assays. Differences across body mass index extended to lung enriched proteins outside the 35. Compartments accounted for 49% of the variance in the 35 protein estimates, against 10% under random reassignment (P < 0.001). At fixed body mass index, type 2 diabetes, hypertension and dyslipidemia showed smaller differences in ARDS biomarkers and were not associated with lower epithelial proteins.

**Conclusions:** Plasma proteins used as biomarkers of inflammation and injury in ARDS vary with body size even in the absence of acute illness. Differences during ARDS may not be attributable to injury alone and may be misinterpreted in patients with obesity due to antecedent variation.

## Introduction

In acute respiratory distress syndrome (ARDS), plasma proteins are used to infer compartment specific patterns of injury (1). Soluble receptor for advanced glycation end products (sRAGE), surfactant protein A2 (SP-A2), surfactant protein D (SP-D) and club cell secretory protein 16 (CC16) are attributed to injury of the alveolar epithelium, angiopoietin-2 and von Willebrand factor (vWF) to endothelial permeability and activation, and interleukin-6 (IL-6), interleukin-8 (IL-8), soluble tumor necrosis factor receptor 1 (sTNFR-1) and the soluble adhesion molecules to the inflammatory response (2). These biomarkers have been used for ARDS prognostication, subphenotype assignment and, more recently, the selection of patients into clinical trials (3–5). However, biomarker concentrations may not fully reflect acute injury when antecedent variation is present.

Body size is one likely source of such variation. Body mass index (BMI) was associated with 44% of the plasma proteins in adults in the United Kingdom and 75% of plasma proteins in adults in China (6, 7). Higher BMI is associated with a greater risk of ARDS development and severity (8, 9). In pooled ARDS Network trials, higher BMI was associated with lower plasma SP-D and higher vWF, suggestive of decreased epithelial and increased endothelial injury in patients with obesity (10). Because body size influences many of these proteins outside disease, the same pattern could be observed in ARDS without any difference in the severity of lung injury. Moreover, type 2 diabetes, hypertension and dyslipidemia are common at higher body size and could account for associations attributed to body size itself.

We measured 35 plasma proteins associated with ARDS risk or mortality in ambulatory adults in the *All of Us* Research Program to define their proteomic patterns across body size before any injury has occurred (11). We aimed to determine whether their associations with body mass index were distinctive across the platform, were reproduced by type 2 diabetes, hypertension and dyslipidemia at fixed body size, and how their differences compared with those reported within ARDS.

## Methods

### Data source

The All of Us Research Program is a national cohort of adults across the United States, with linked surveys, physical measurements and electronic health records (11). We used the Controlled Tier curated data repository, version C2025Q4R6, with a data cutoff of January 1, 2025. Participants provided written informed consent under a protocol approved by the All of Us Research Program institutional review board. The Mass General Brigham institutional review board classified this secondary analysis of deidentified data as not human subjects’ research.

### Selected proteins and measurement

We registered the selected proteins on the Open Science Framework before any analysis (https://doi.org/10.17605/OSF.IO/9YEHP). Our study included 35 proteins associated with ARDS development or mortality from a systematic review (2). The proteins were organized into pathophysiological compartments that included epithelial (4 proteins), endothelial permeability (6 proteins), endothelial activation and coagulation (5 proteins), inflammation (17 proteins) and myocardial (3 proteins).

Plasma proteins were measured by proximity extension assay on the Olink Explore HT platform, which reports relative concentrations as normalized protein expression (NPX) values on a log base 2 scale (12, 13). The release contained 5,420 protein assays for 9,969 participants, covering 5,416 proteins. We applied a between-person standard deviation of at least 0.4 NPX, because an assay with small variance may not show a difference by body size. Two proteins fell just below this threshold and were retained, with a sensitivity analysis excluding them in the online supplement.

### Analytic cohort

Proteomic samples were drawn from the 14,521 participants with long-read whole genome sequencing, selected to reflect the range of genetic ancestry in the program. The analytic cohort was participants with a recorded plasma collection date, a recorded BMI, age 18 years or older, available genetic principal components, and no inpatient encounter spanning the collection date. We excluded samples failing quality control (online supplement). Comparisons are in **Table E1**.

### Exposure and covariates

Our exposure was BMI defined from physical measurements where available and otherwise from the health record nearest to the collection date.

We adjusted for age at plasma collection as a restricted cubic spline with four degrees of freedom, sex at birth, ever and current smoking, thirteen chronic conditions, and the first ten genetic principal components, included because protein measurements on this platform are influenced by common genetic variants whose frequencies differ across ancestries. We imputed missing values for sex at birth and smoking with 20 draws combined by Rubin’s rules (14). We defined chronic conditions from diagnoses in the electronic health record and required two occurrences on distinct dates before plasma collection (15, 16). The diagnosis codes are available in **Table E2**. We treated type 2 diabetes, hypertension and dyslipidemia as metabolic conditions. We defined a metabolically healthy group as participants with at least one diagnosis recorded in the health record but no recorded diagnosis of a metabolic condition. We followed the RECORD extension for studies using routinely collected health data (17).

### Statistical analysis

We standardized each protein assay to zero mean and unit variance given that NPX is an arbitrary unit whose distribution differs between assays. We modeled BMI with a restricted cubic spline with three degrees of freedom, fitted by ordinary least squares (18). We reported every BMI to protein association as the difference in standardized concentration between a BMI of 35 and 25 kg/m^2^, a linear combination of the three spline coefficients with a 95% confidence interval from its variance.

We repeated the same model for all 5,420 assays in the *All of Us* platform, which provided a distribution of body size associations across the entire proteome. We then reported the position of each protein in the platform as the proportion of assays with an estimate for BMI at or below its own.

### Secondary analyses

To test whether the 35 estimates were organized by pathophysiological compartment, we calculated the proportion of variance lying between compartments rather than within, against 10,000 permutations of the set labels, and contrasted each compartment with the rest by a two-sided permutation test. We then modeled type 2 diabetes, hypertension and dyslipidemia each as the exposure in place of BMI, holding BMI fixed with the same spline and adjusting for the ten remaining conditions, with the mutually adjusted version in **Table E3**, and tested whether their estimates agreed as closely as a single shared pattern would produce. We defined tissue specificity from the Human Protein Atlas version 25.1 (19), and assigned each protein’s cell type from the single cell cluster in which it was most highly expressed. We compared mean estimates by how broadly each protein is expressed, recomputed each protein’s position within its own group, and repeated the compartment variance analysis after centering each estimate within its tissue specificity class. Finally, we tested departure from a straight line by an F test against a model linear in BMI, and differences across genetically inferred ancestry groups by a joint F test on the interaction terms.

In sensitivity analyses, we adjusted for creatinine and medication exposure, replaced BMI with waist circumference and waist to hip ratio, excluded participants admitted to hospital within 30 days of collection, restricted to participants with any recorded diagnosis and to genetically unrelated participants, and refitted with robust standard errors (20) and assay plate. Knot positions for spline models, classification thresholds and full specifications are in the online supplement.

We performed analyses using R version 4.4 and followed the STROBE recommendations for cross sectional studies (21), with the completed checklist in **Table E9**.

## Results

Of 9,969 participants with plasma proteomic measurements in *All of Us*, 8,680 met our criteria (**Figure 1**, **Table 1**). Median age was 50 years, 50% were female, median BMI was 27.2 kg/m^2^, and 30% were of European genetic ancestry. The 35 to 25 kg/m^2^ contrast spanned the 34th to 86th percentile of BMI in this cohort.

**Figure 1.**
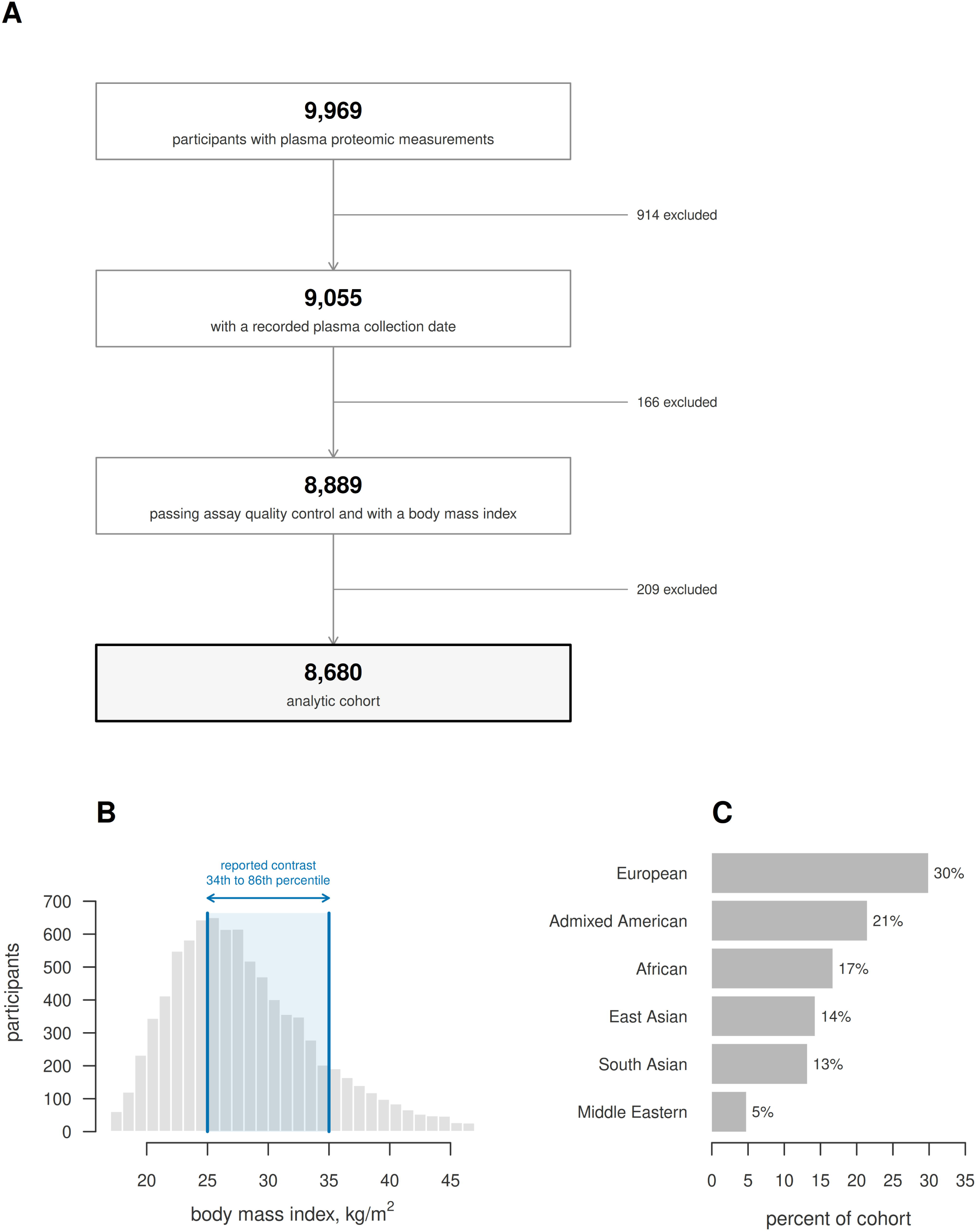
The study cohort. A, participants in the *All of Us* proteomics release and the criteria applied to reach the analytic cohort. B, the distribution of body mass index, with the reported contrast between 25 and 35 kg/m^2^ shaded, spanning the 34th to the 86th percentile. Participants with a body mass index below 17 kg/m^2^ (n = 28) or at or above 47 kg/m^2^ (n = 166) are not shown. C, genetically inferred ancestry as a percentage of the cohort.

**Table 1.** Characteristics of 8,680 adults in the *All of Us* Research Program.

| Characteristic, median (interquartile range) or n (%) | Value |
| --- | --- |
| Participants, n | 8,680 |
| Age, years | 50 (33 to 63) |
| Sex at birth |  |
| Female | 4,309 (49.6) |
| Male | 3,901 (44.9) |
| Not recorded | 470 (5.4) |
| Body mass index, kg/m<sup>2</sup> | 27.2 (23.8 to 31.8) |
| Body mass index category |  |
| <25 | 2,910 (33.5) |
| 25 to <30 | 2,897 (33.4) |
| 30 to <35 | 1,613 (18.6) |
| ≥35 | 1,260 (14.5) |
| Waist circumference, cm | 92.0 (81.5 to 103.5) |
| Measured in | 7,848 (90.4) |
| Smoking |  |
| Ever | 2,924 (33.7) |
| Current | 1,321 (15.2) |
| Not reported | 321 (3.7) |
| Creatinine, mg/dL | 0.85 (0.71 to 1.00) |
| Measured in | 3,973 (45.8) |
| eGFR, mL/min/1.73 m<sup>2</sup> | 93 (77 to 107) |
| Measured in, n | 3,770 (43.4) |
| Genetically inferred ancestry |  |
| European | 2,591 (29.9) |
| American | 1,859 (21.4) |
| African | 1,445 (16.6) |
| East Asian | 1,235 (14.2) |
| South Asian | 1,140 (13.1) |
| Middle Eastern | 410 (4.7) |
| Admitted to hospital within 30 days of collection | 250 (2.9) |
| At least one diagnosis record | 7,232 (83.3) |
| Metabolic conditions, two occurrences |  |
| Type 2 diabetes | 916 (10.6) |
| Hypertension | 1,998 (23.0) |
| Dyslipidemia | 1,964 (22.6) |
| Other chronic conditions |  |
| Cancer | 756 (8.7) |
| Asthma | 690 (7.9) |
| Obstructive sleep apnea | 544 (6.3) |
| Coronary artery disease | 476 (5.5) |
| Heart failure | 259 (3.0) |
| Chronic obstructive pulmonary disease | 243 (2.8) |
| Stroke | 242 (2.8) |
| Atrial fibrillation | 237 (2.7) |
| Alcohol use disorder | 230 (2.6) |
| Interstitial lung disease | 68 (0.8) |
| Metabolically healthy stratum |  |
| No occurrence of the three conditions | 5,331 (61.4) |
| With at least one diagnosis record | 3,883 (44.7) |
| Type 2 diabetes, any occurrence | 1,132 (13.0) |
| Hypertension, any occurrence | 2,455 (28.3) |
| Dyslipidemia, any occurrence | 2,425 (27.9) |
| Medications |  |
| Statin in the year before collection | 1,076 (12.4) |
| Metformin in the year before collection | 464 (5.3) |

Values are median (interquartile range) or n (%), with percentages of all 8,680 participants.
Rows labeled measured in give the number for whom the measurement was available. Chronic conditions required two diagnosis codes on distinct dates before plasma collection, and the any occurrence rows beneath the metabolically healthy stratum use a single code. The metabolically healthy stratum required no recorded occurrence of type 2 diabetes, hypertension or dyslipidemia before plasma collection, a more stringent exclusion, and the single-occurrence prevalences it rests on are shown beneath it. Estimated glomerular filtration rate was calculated with the 2021 CKD-EPI creatinine equation, which does not include a race coefficient. Creatinine was not measured in every participant and does not enter the primary models.

Across the 5,420 protein assays on the platform, 4,296 associations with body size were positive (79.3%), with a median of 0.065 SD (**Figure 2**, **Table E4**). All four epithelial proteins were lower at higher BMI, each below at least 92% of assays. sRAGE was 0.350 SD lower (95% CI 0.297 to 0.404) and below 98.8% of assays, SP-A2 was 0.084 SD lower (95% CI 0.027 to 0.140), SP-D was 0.083 SD lower (95% CI 0.027 to 0.138) and CC16 was 0.081 SD lower (95% CI 0.029 to 0.133). IL-6 and E-selectin were 0.534 SD (95% CI 0.481 to 0.588) and 0.465 SD (95% CI 0.411 to 0.520) higher, above 99.5% and 99.2% of assays on the platform, respectively.

**Figure 2.**
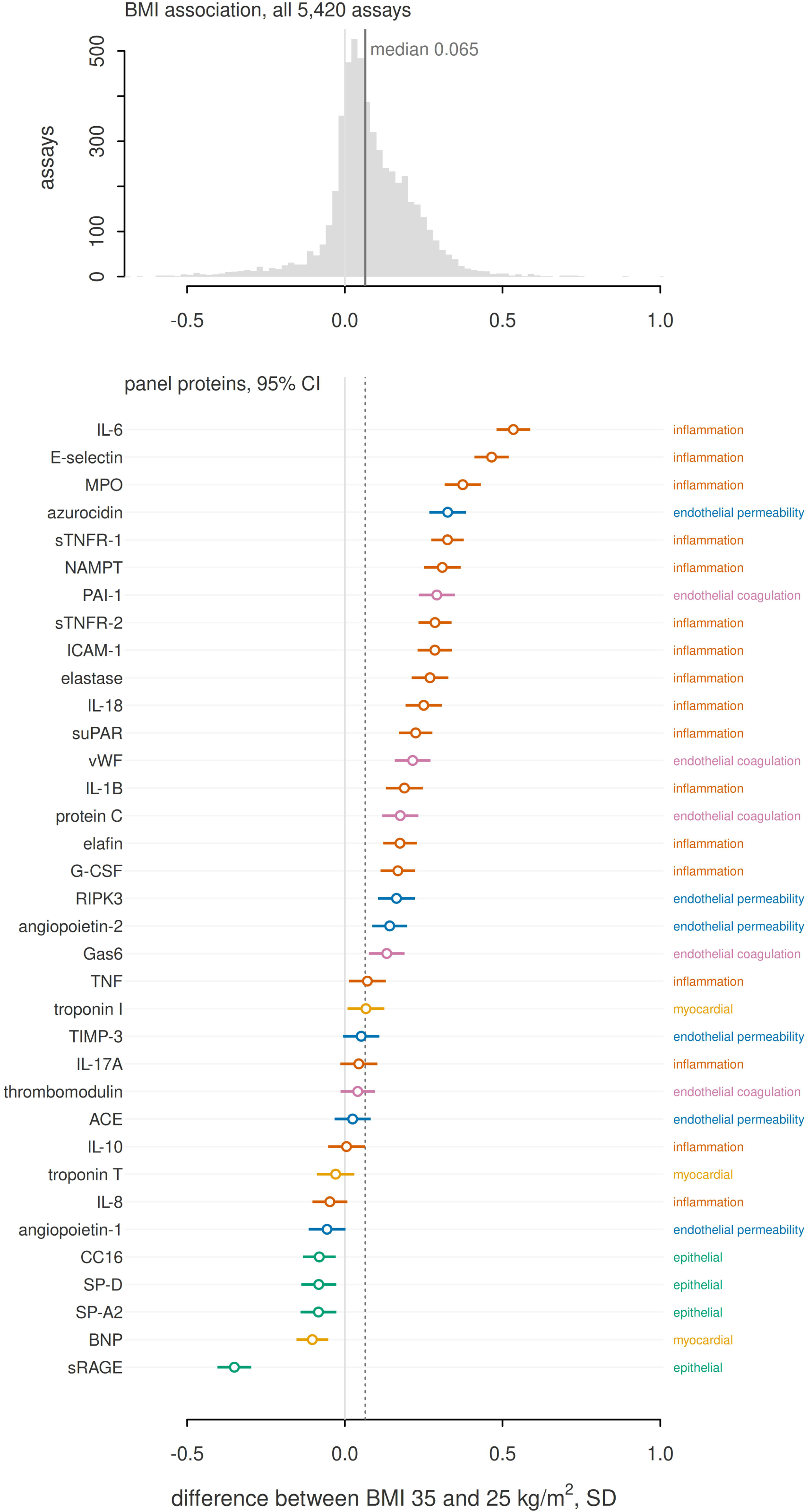
Body mass index associations across the platform and within the 35 ARDS proteins. Upper, the distribution of the difference between a body mass index of 35 and 25 kg/m^2^, in standard deviations, across all 5,420 assays on the Olink Explore HT platform. Lower, each of the 35 proteins associated with ARDS with its 95% confidence interval on the same axis, ordered by estimate, colored and labeled by compartment. The five compartments are epithelial, endothelial permeability, endothelial activation and coagulation, inflammation and myocardial. A light vertical line marks zero in both panels. The platform median is shown as a labeled line in the upper panel and a dotted line in the lower.

The difference per unit of BMI was not constant across the range for 25 of the 35 proteins. For three of the four epithelial proteins the concentration fell more steeply at lower BMI than at higher BMI, with fitted relationships and 95% confidence bands shown in **Figure 3**. sRAGE was 0.295 SD higher at a BMI of 20 than at 25 (95% CI 0.238 to 0.353) and 0.434 SD lower at 40 than at 25 (0.372 to 0.495), falling about three times as steeply in the lowest third of the BMI range as in the highest. SP-D and CC16 fell in the same pattern and SP-A2 at a nearly constant rate. Slopes across the BMI range, tests for departure from linearity and a shape classification for each protein are in **Table E5**.

**Figure 3.**
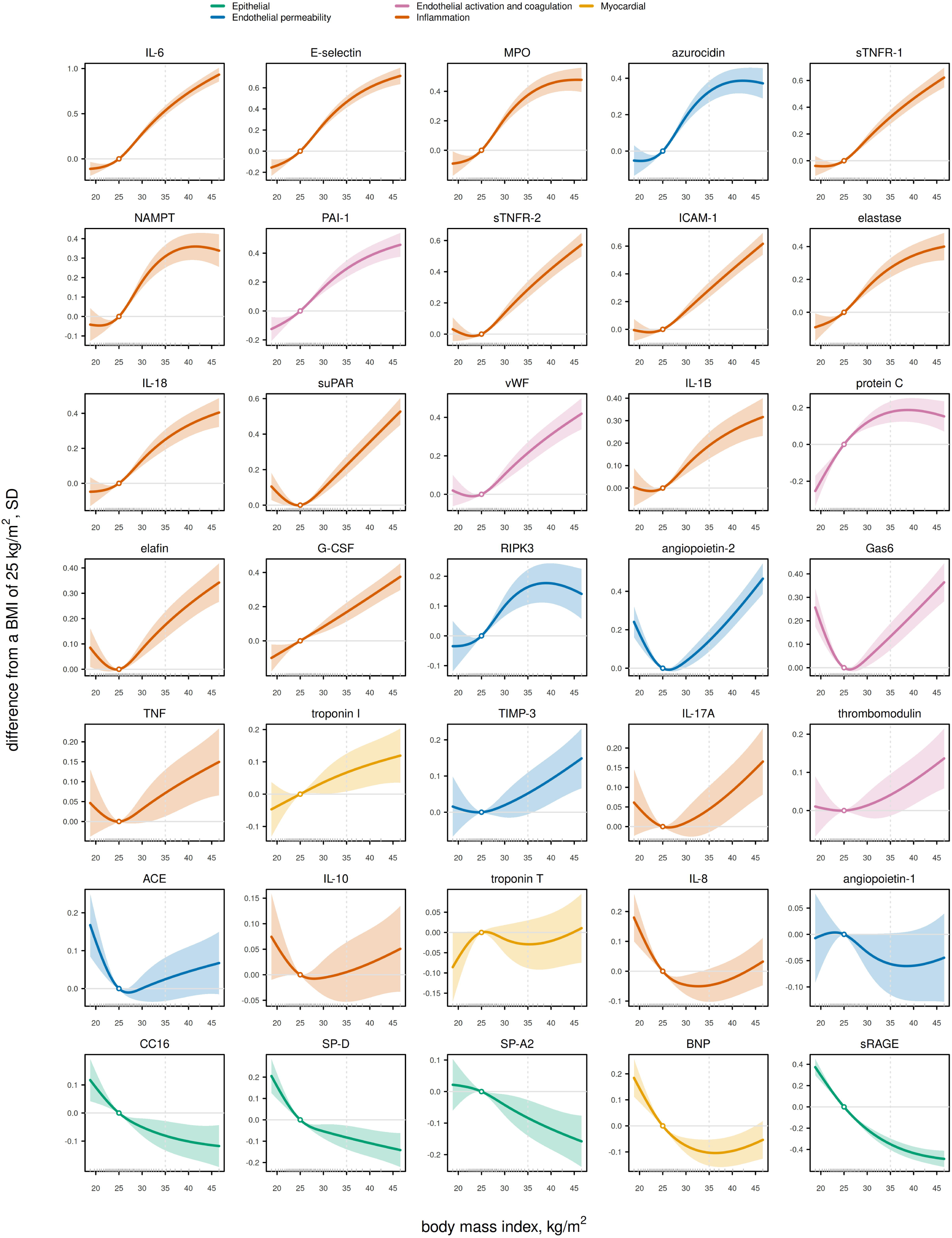
Fitted associations between body mass index and each ARDS protein. Curves show the difference in standardized concentration from a body mass index of 25 kg/m^2^, with 95% confidence bands. The estimate and its interval are zero at 25 by construction, marked by the open point, and the band widens with distance from that reference. Tick marks along the horizontal axis show the distribution of body mass index in the cohort, at every second percentile. The dotted vertical line is a body mass index of 35, the upper end of the reported contrast. Colors denote compartment. Vertical scales differ between panels.

Proteins in the same compartment had similar associations with body size. Compartment membership accounted for 49% of the variance in the 35 protein estimates, against 10% when proteins were randomly reassigned (permutation P < 0.001). The epithelial set averaged 0.323 SD below the rest of the 35 (95% CI −0.522 to −0.124) and the inflammation set 0.184 SD above it (0.074 to 0.294), both by permutation (P < 0.001 and P = 0.002). Across the platform, proteins classified in the Human Protein Atlas as made mainly in one or a few tissues had less positive associations with body size than proteins made across many tissues, 0.060 SD against 0.113 SD, a difference of 0.053 SD (95% CI 0.045 to 0.060). Levels of epithelial proteins were lower even against that background. Among proteins made mainly in a single tissue, sRAGE still ranked below 96% of assays and SP-A2, SP-D and CC16 below 83%. Compartment membership still accounted for 44% of the variance among the 35 estimates after we centered each estimate within its tissue specificity class, against 10% expected (P < 0.001).

Associations with BMI extended to lung epithelial proteins outside the 35 we selected. Nine proteins on the *All of Us* platform are produced mainly in lung tissue. The six proteins whose predominant cell type is alveolar, ciliated or secretory occupied the six lowest positions, the lowest possible rank sum (exact P = 0.012). SCGB3A2, a secretoglobin related to CC16, had the most negative association, 0.498 SD lower (95% CI 0.446 to 0.551) and lower than 99.7% of assays on the platform. The three proteins that were not epithelial in origin, RTKN2 and the macrophage markers, MSR1 and CCL18, were higher at higher BMI, by 0.027 to 0.403 SD.

At fixed BMI, type 2 diabetes, hypertension and dyslipidemia were associated with smaller differences in the plasma proteome than BMI (**Figure 4**). Across the 5,420 assays, the median difference was 0.038 SD for type 2 diabetes, 0.013 for hypertension and 0.016 for dyslipidemia, against 0.065 SD for body size. These conditions did not associate with lower epithelial proteins. In contrast, epithelial proteins were even lower in participants who were classified as metabolically healthy (**Table E8**). Metabolic conditions did not yield the same pattern across the 35 proteins. Their estimates for the 35 proteins correlated with one another by a mean rank correlation of 0.458. Had the three shared one underlying effect, their estimates would have differed only by sampling error, which would give a mean correlation of 0.794 (95% simulation interval 0.712 to 0.866). The observed value of 0.458 was lower than all 10,000 simulated values (P < 0.001).

**Figure 4.**
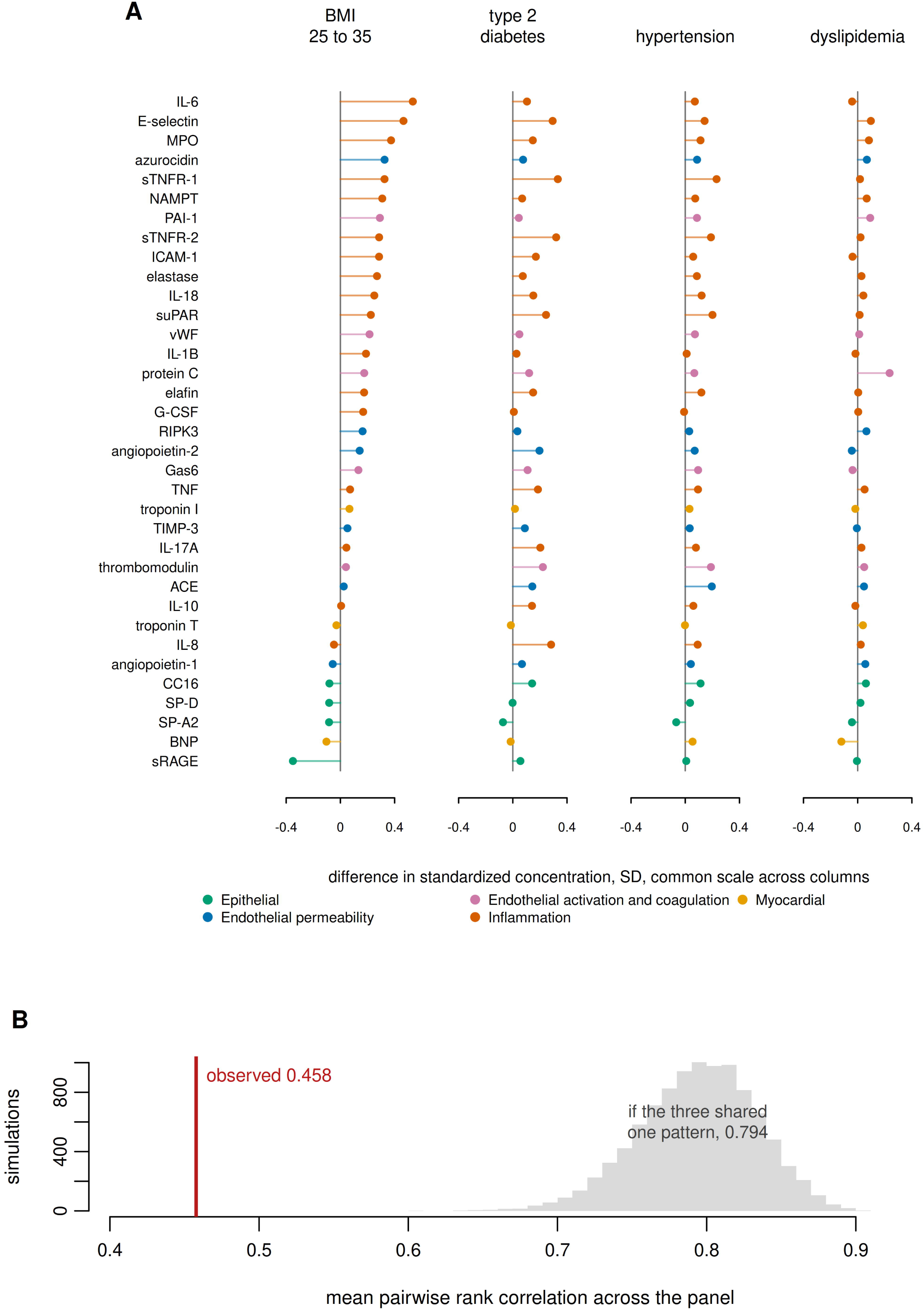
ARDS proteins under body mass index and the three metabolic conditions. A, the difference in standardized concentration for each of the 35 proteins under four exposures, ordered by the body mass index estimate and colored by compartment. Type 2 diabetes, hypertension and dyslipidemia were each modeled at fixed body mass index. The vertical line in each column marks zero and points to its right are positive. The four columns share one scale. Green points are the epithelial proteins, the only compartment whose direction changes between body mass index and the metabolic conditions. B, the mean pairwise rank correlation among the three conditions across the 35 proteins, against the distribution expected if the three shared a single underlying pattern.

BMI to protein associations varied across the six genetically inferred ancestry groups. Among the epithelial proteins, the variation was confined to CC16 (interaction P < 0.001) where it was higher with BMI in participants of African ancestry and lower with BMI in the other five ancestry groups. Group specific estimates and Cochran’s Q are in **Table E6**.

The estimates were stable across alternative specifications (**Table E7)**. Adjusting for creatinine changed the median estimate by 0.001 SD. Substituting waist circumference for BMI reproduced the proteomic pattern with a correlation of 0.978, and 0.93 for waist to hip ratio. Robust standard errors, adjustment for assay plate, and exclusion of related participants each changed no estimate by more than 0.023 SD.

Lastly, in comparison to estimates in ARDS, three of the five ARDS proteins with a reported within ARDS trend agreed in direction with estimates in *All of Us* (**Table 2**). SP-D was lower, and vWF and protein C were higher. IL-8 decreased with BMI in ARDS and was in the same direction in *All of Us*, although the confidence interval included zero. Discordance took two forms. sTNFR-1, ICAM-1 and PAI-1 were unrelated to BMI in ARDS while being among the strongest positive associations in *All of Us*. Only IL-6 was opposite in direction, lower in ARDS and the largest positive association with BMI in All of Us.

**Table 2.**
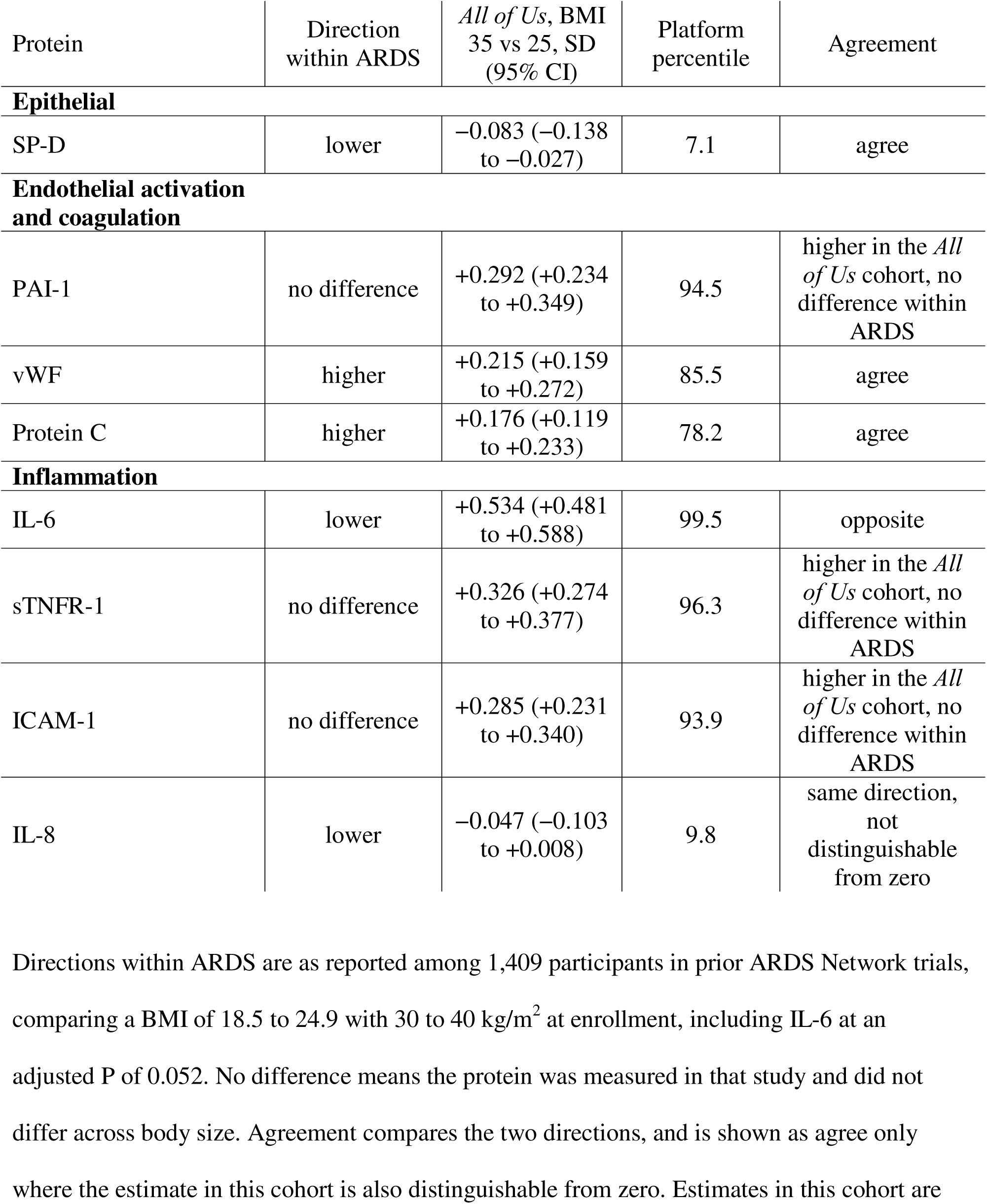
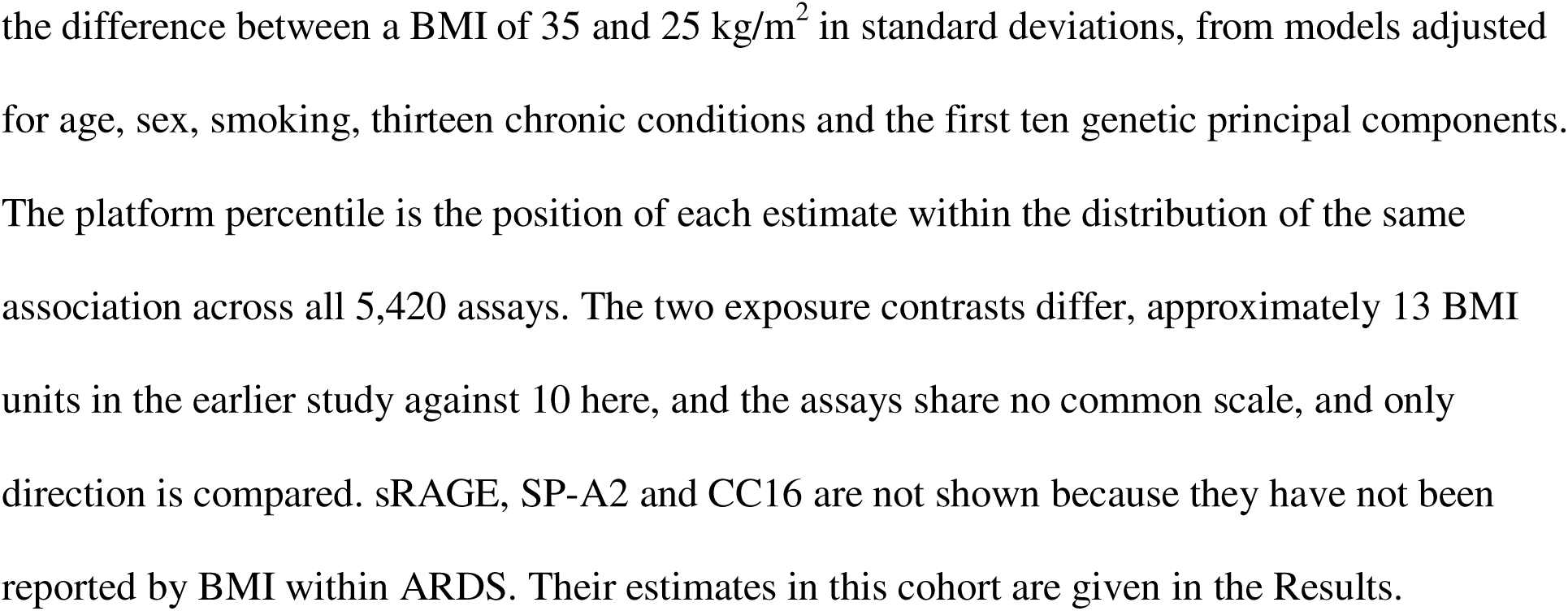
Direction of the body size associations in the *All of Us* Research Program and within published studies of ARDS.

| Protein | Direction within ARDS | *All of Us*, BMI 35 vs 25, SD (95% CI) | Platform percentile | Agreement |
| --- | --- | --- | --- | --- |
| **Epithelial** |  |  |  |  |
| SP-D | lower | −0.083 (−0.138 to −0.027) | 7.1 | agree |
| **Endothelial activation and coagulation** |  |  |  |  |
| PAI-1 | no difference | +0.292 (+0.234 to +0.349) | 94.5 | higher in the *All of Us* cohort, no difference within ARDS |
| vWF | higher | +0.215 (+0.159 to +0.272) | 85.5 | agree |
| Protein C | higher | +0.176 (+0.119 to +0.233) | 78.2 | agree |
| **Inflammation** |  |  |  |  |
| IL-6 | lower | +0.534 (+0.481 to +0.588) | 99.5 | opposite |
| sTNFR-1 | no difference | +0.326 (+0.274 to +0.377) | 96.3 | higher in the *All of Us* cohort, no difference within ARDS |
| ICAM-1 | no difference | +0.285 (+0.231 to +0.340) | 93.9 | higher in the *All of Us* cohort, no difference within ARDS |
| IL-8 | lower | −0.047 (−0.103 to +0.008) | 9.8 | same direction, not distinguishable from zero |

Directions within ARDS are as reported among 1,409 participants in prior ARDS Network trials, comparing a BMI of 18.5 to 24.9 with 30 to 40 kg/m<sup>2</sup> at enrollment, including IL-6 at an adjusted P of 0.052. No difference means the protein was measured in that study and did not differ across body size. Agreement compares the two directions, and is shown as agree only where the estimate in this cohort is also distinguishable from zero. Estimates in this cohort are
the difference between a BMI of 35 and 25 kg/m<sup>2</sup> in standard deviations, from models adjusted for age, sex, smoking, thirteen chronic conditions and the first ten genetic principal components. The platform percentile is the position of each estimate within the distribution of the same association across all 5,420 assays. The two exposure contrasts differ, approximately 13 BMI units in the earlier study against 10 here, and the assays share no common scale, and only direction is compared. sRAGE, SP-A2 and CC16 are not shown because they have not been reported by BMI within ARDS. Their estimates in this cohort are given in the Results.

## Discussion

In 8,680 ambulatory adults, the plasma proteins associated with ARDS risk and mortality varied with body size in the absence of acute illness. All four proteins related to epithelial injury were lower at higher BMI, while the inflammatory proteins were higher as a set. Each epithelial protein was below at least 92% of the 5,420 assays measured in *All of Us*, and IL-6 and E-selectin above 99%. Compartment membership accounted for nearly 50% of the variance among the 35 estimates. At fixed body size, type 2 diabetes, hypertension and dyslipidemia were not associated with lower epithelial proteins.

The four epithelial proteins were lower at higher body size, and their mean estimate was the lowest of the five compartments. These proteins are produced from different epithelial cells in the lung. sRAGE is released by type I alveolar cells, SP-A2 and SP-D by type II alveolar cells, and CC16 by club cells of the distal airway. Data from the Human Protein Atlas extend our results beyond the 35 we selected. Of the nine proteins produced primarily in lung tissue, the six of epithelial origin had the most negative associations with BMI, including SCGB3A2, a small, secreted protein of airway epithelium related to CC16. Of the three proteins that were not epithelial, two are expressed mainly by macrophages (MSR1, CCL18) and were higher at higher BMI. Proteomic differences across body size appear confined to proteins produced in the lung epithelium.

The lung, however, does not appear to produce less of these proteins. Per-cell expression of the protein transcripts from donors without lung disease did not differ across BMI (22). Levels of SP-A2, SP-D and CC16 were also similar across BMI in bronchoalveolar lavage fluid from adults undergoing non-thoracic surgery (23). In the same participants, the alveolar lipidome, including surfactant phospholipids, also showed no significant variation by body weight (24). Neither transcription in lung cells nor protein concentration in bronchoalveolar fluid tracks with the lower plasma values we observed. CC16 is a partial exception. The proportion of CC16 expressing cells is lower in the small airways of mice and humans with obesity (25), and lower serum CC16 accompanies club cell loss in smokers (26). Fewer club cells could account for the lower CC16 we observed, and possibly for SCGB3A2.

Lower lung epithelial protein concentrations at higher BMI likely arise from mechanisms other than production. Excess adiposity collapses dependent lung regions and may reduce the amount of aerated alveolar surface needed for these proteins to enter the blood (27, 28). Plasma concentrations are also affected by permeability of the alveolar-capillary barrier (26), although it is unknown whether adiposity influences permeability. Clearance and dilution are less likely mechanisms based on our findings. Adjusting for serum creatinine augmented differences across BMI in the lung epithelial proteins. And while plasma volume is known to increase with body size (29, 30), dilution does not fit with our findings of a predominantly positive association between BMI and the plasma proteome.

Not accounting for antecedent variation across body size may lead to misinterpretation of ARDS biomarkers. A threshold fixed in one body size distribution may not identify the same characteristics at another. The difference is systematic with a predictable direction. Prior to ARDS, patients with obesity already show lower epithelial and higher endothelial and inflammatory markers. Measurements read in isolation would attribute these patterns to injury, whether they are used for ARDS diagnosis, subphenotype assignment, or clinical trial selection. IL-6 was the only protein whose direction disagreed with the trend reported within ARDS, higher with BMI in *All of Us* and lower in the pooled ARDS data. Patients with obesity may meet clinical criteria for ARDS by alveolar compression from excess adiposity, which produces hypoxemia and opacities often indistinguishable from those of inflammatory injury (31, 32), and would then have lower IL-6 at the same degree of hypoxemia. Obesity may also blunt the acute inflammatory response. Separating these requires measurements in the same people before and during illness.

Obesity co-exists with type 2 diabetes, hypertension and dyslipidemia, which makes it important to disentangle their separate associations with the plasma proteome. At fixed BMI, these other metabolic conditions had smaller associations across the platform than BMI, did not separately lower the epithelial proteins in the lung, and their estimates across the 35 proteins were less correlated than a single shared effect would yield. Epithelial proteins were even lower still across BMI in adults without metabolic disease. Mass loading on the lung and alveolar collapse may contribute more to the lower epithelial proteins than a purely metabolic driver. Adjusting for the three conditions as a single exposure would obscure that separation.

Several limitations impact our interpretations. The comparison of our estimates to ARDS cohorts is between studies and directional only as those trials report immunoassay concentrations instead of relative values. Earlier cohorts measured by immunoassay have reported SP-D and CC16 lower at higher BMI, which argues against a platform artifact (25, 33). The design is cross sectional and describes variation across individuals, not change within them, and cannot establish direction (33). We cannot establish what fraction of the within ARDS difference is antecedent, and differences related to injury may be superimposed on it. Confounding by unmeasured factors that track with body size cannot be excluded. *All of Us* enrolls volunteers rather than a probability sample. Associations differed across ancestry groups, which cautions against transferring estimates between populations.

## Conclusion

Plasma proteins are used as biomarkers of inflammation and injury in ARDS and vary with body size in the absence of acute illness. Differences in these proteins during ARDS may not be specific to injury and require consideration of antecedent variation for their interpretation.

## Acknowledgments

We gratefully acknowledge *All of Us* participants for their contributions, without whom this research would not have been possible. We also thank the National Institutes of Health’s *All of Us* Research Program for making available the participant data examined in this study.

## Funding

This work was supported by the National Heart, Lung, and Blood Institute of the National Institutes of Health under award number 5K23HL175128. The funder had no role in the design or conduct of the study, in the analysis or interpretation of the data, or in the preparation of the manuscript.

## Conflicts of Interest

The authors have no conflicts of interest to disclose. No author or their institution received payments or services from a third party for any aspect of the submitted work in the past 36 months.

## Data Availability

This study used data from the *All of Us* Research Program’s Controlled Tier Dataset C2025Q4R6, available to authorized users on the Researcher Workbench. Individual level data cannot be redistributed under the program’s Data Use and Registration Agreement. Analysis code and the estimates for all 5,420 assays are available from the corresponding author on reasonable request, for use within the Researcher Workbench by authorized users.

## Author contributions

T.G. conceived the study, designed and performed the analysis, and wrote the manuscript. L.B. contributed to the design and to the interpretation of the findings and revised the manuscript. M.C. supervised the study, contributed to its design and interpretation, and revised the manuscript. All authors approved the final version and agreed to be accountable for all aspects of the work.

## Funding

Supported by the National Heart, Lung, and Blood Institute of the National Institutes of Health under award number 5K23HL175128. The content is solely the responsibility of the authors and does not necessarily represent the official views of the National Institutes of Health.

## Conflicts of interest

The authors have no conflicts of interest to disclose. No author or their institution received payments or services from a third party for any aspect of the submitted work.

## Registration

The 35 proteins and their five compartment sets were registered on the Open Science Framework before any analysis (https://doi.org/10.17605/OSF.IO/9YEHP).

## Notes

### Competing Interest Statement

The authors have declared no competing interest.

### Clinical Protocols

https://doi.org/10.17605/OSF.IO/9YEHP

### Author Declarations

The Institutional Review Board of Mass General Brigham waived ethical approval for this work, having determined that this secondary analysis of de-identified data is not human subjects research. The Institutional Review Board of the National Institutes of Health All of Us Research Program gave ethical approval for the protocol under which participants provided written informed consent.

