## Supplementary material for "Plasma Biomarkers of Acute Respiratory Distress Syndrome Vary with Body Size in the Absence of Acute Illness": Online Data Supplement

### E1. Panel definition and registered criteria

Set membership was registered on the Open Science Framework before any analysis. The set registered as alveolar epithelial is named epithelial in the manuscript, with membership unchanged, because CC16 is secreted by club cells of the distal airway. A protein belongs to a set if van der Zee and colleagues assigned it that role in their Table 3 or Table 4, plasma compartment only. The rule was chosen so that membership is reproducible by a third party and independent of any result in these data, and it was not revised after the estimates were seen.

Membership additionally required presence on the Olink Explore HT panel and a between-person standard deviation of at least 0.4 NPX in the population to which the sets are applied. Two proteins fell below that threshold in this cohort, protein C at 0.372 and Gas6 at 0.388, both in the endothelial activation and coagulation set, and both were retained. The criterion was registered to exclude assays without measurable signal, and neither protein is of that kind. Protein C has an association of 0.176 standard deviations with an interval clear of zero, and both are abundant plasma proteins under tight physiological control, for which modest between-person variation is expected rather than a sign of assay failure. Excluding both leaves the compartment result unchanged. Among the remaining 33 estimates, 49.4% of the variance lay between sets against 49% for the full panel, with a permutation P value of 0.0004.

Seven proteins assigned a role in the source review are not on the Olink Explore HT panel and could not be included: surfactant protein A1, insulin-like growth factor 1, inhibin beta A, C-reactive protein, TREM1, HMGB1 and adiponectin. C-reactive protein is the most frequently studied marker in that review, and its absence bounds what any analysis using these sets can say about the inflammatory compartment.

Two discrepancies with other sources were declared at registration and are reported here rather than resolved. ICAM-1 and E-selectin are labeled pro-inflammatory by van der Zee and endothelial by the American Thoracic Society workshop report. The registered rule follows van der Zee, so both sit in the inflammation set. Because both carry large positive associations with body size, the alternative assignment changes which of the two positive sets leads. Endothelial activation and coagulation would average 0.230 SD and inflammation 0.212, leaving the two indistinguishable, where under the registered rule inflammation is clearly the higher. The epithelial set is unaffected either way, remaining 0.149 SD below zero and the lowest of the five, so the finding the paper rests on does not depend on the assignment. Angiopoietin-1 enters through the angiopoietin-1 to angiopoietin-2 ratio in Table 4 of the source. MUC1 and TEK have no role assignment in the source and were excluded.

### E2. Concept identifiers

Concept identifiers for every variable drawn from the electronic health record, with the Observational Medical Outcomes Partnership concept and the descendant expansion used, are listed in Table E2.

### E3. Selection of participants for proteomics

The program reports that proteomics and RNA sequencing samples were drawn from participants who already had long-read whole genome sequencing, and were generally chosen to reflect the range of genetic ancestry in the program. Long-read participants were themselves selected by each genome center under its own criteria, all requiring existing short-read sequencing and sufficient high molecular weight DNA at the biobank, with record availability used as a prioritization factor. One center selected participants who identified as African American or Black, two selected participants who identified as Hispanic, one selected on computed genetic ancestry with an emphasis on admixed individuals and family groups, and one included participants with a known disease expected to benefit from long-read sequencing.

Almost all assayed participants came from one center. We describe how the assayed differed from the rest of the long-read cohort, and report logistic models for what predicted being assayed, in Table E1. Across the long-read manifest as a whole, the proportion assayed fell with body size, from 75% in the lowest tenth of BMI to 42% in the highest, and the assayed had a median BMI of 27.1 kg/m2 against 30.2 in the rest of the manifest. That gradient was between centers rather than within them. The centers whose participants were almost never assayed applied different selection criteria and differed in body size from the center that supplied the cohort. Within the center that supplied the cohort, the proportion assayed ranged from 91.8% to 94.3% across tenths of the BMI distribution with no order to it, and body size did not predict selection.

### E4. Sample quality control

For each sample we computed the proportion of assays below the limit of detection and the minimum across assays of that sample’s value expressed in standard deviations of that assay’s distribution across samples. We excluded samples exceeding 5% on the first or falling below minus 20 on the second. Fewer than 20 samples met at least one criterion, and both criteria identified the same samples.

### E5. Missing covariates and multiple imputation

Sex at birth was not recorded for 470 participants and 321 did not answer the smoking items. We retained them and imputed by multiple imputation with 20 draws, fitting logistic imputation models containing the BMI and age splines, ancestry, the conditions, the principal components and the 35 panel proteins, drawing the imputation model coefficients from their estimated sampling distribution at each draw, and combining estimates across the 20 draws by Rubin’s rules, which add the variation between draws to the variation within them. We set current smoking to zero in any draw in which ever smoking was imputed as zero. Principal components and inferred ancestry are those the program provides from short read whole genome sequencing.

### E6. Chronic conditions and the metabolically healthy stratum

We defined chronic conditions from diagnoses in the electronic health record, using Observational Medical Outcomes Partnership concepts and every more specific concept beneath them, requiring two occurrences on distinct dates before collection. The thirteen were hypertension, dyslipidemia, type 2 diabetes, cancer, asthma, obstructive sleep apnea, coronary artery disease, heart failure, chronic obstructive pulmonary disease, cerebrovascular disease, atrial fibrillation, disorder caused by alcohol and interstitial lung disease. Conditions are named by the concept queried, which is broader than the colloquial term for two of them. Requiring two occurrences avoids counting a diagnosis recorded once to justify a test or recorded in error.

The two definitions are deliberately asymmetric. We count a participant as having a condition only after two occurrences, which avoids calling someone diseased on the strength of one code, but a single occurrence is enough to remove a participant from the metabolically healthy stratum, which is meant to contain only participants in whom the conditions can be excluded. Applying the two occurrence rule to the stratum would admit a further 561 participants, with a median BMI of 28.2 kg/m2 against 26.2 and a median age of 53 years against 40, most of them carrying a single code for hypertension or dyslipidemia.

Participants with no diagnosis records are classified as free of every condition by construction rather than by observation. We addressed this in two ways, by restricting the metabolically healthy stratum to participants with at least one diagnosis record, which we report as the primary analysis, and by treating the condition status of participants without records as missing and imputing it. That imputation assumes their condition prevalence resembles that of participants with records given the covariates, which is unlikely to hold exactly. We read the two analyses as bounds rather than as a correction.

### E7. Creatinine

We took the value nearest to and before collection within two years and entered it on the log scale. We carried it as a sensitivity analysis under the same imputation framework, imputing log creatinine by linear regression, and report the complete case version alongside.

Under complete case analysis, restricted to the participants in whom creatinine was measured, the largest change from the full cohort estimate was 0.080 SD. Within those same participants, adding creatinine to the model moved no estimate by more than 0.028 SD, with a median of 0.003, and the change therefore reflected which participants had a creatinine measured rather than the effect of adjusting for it. The fraction of missing information for the imputed covariates was below 0.004 for every protein.

### E8. Estimation and the platform reference distribution

We modeled BMI with a restricted cubic spline with three degrees of freedom, placing the interior knots at the 33rd and 67th percentiles and the boundary knots at the first and 99th. The reported association is a linear combination of the three spline coefficients, with weights equal to the difference in the spline basis at a BMI of 35 and at 25, and a standard error from the model covariance for that combination. Because the weights are zero on every other column, no other term contributes.

We recomputed the standard error for every assay with the HC3 estimator, which does not assume that the scatter of participants around the fitted curve is the same at every BMI. Samples were assayed in 61 projects across two batches, and we repeated the panel analysis with plate entered as a fixed effect.

### E9. Organization by compartment

We compared the proportion of variance lying between the five sets with its distribution under 10,000 random permutations of the set labels, taking the P value as one plus the number of permutations at or above the observed value, divided by one plus the number of permutations. We repeated this weighting each estimate by the inverse of its variance. We contrasted each set with the remainder of the panel as a difference in means, with a two sided permutation P value, because a set may contain as few as three proteins.

### E10. Metabolic conditions at fixed body size

We entered type 2 diabetes, hypertension and dyslipidemia each as an exposure with BMI held fixed by the same spline, adjusting for the ten non metabolic conditions and not for one another, since mutual adjustment removes the shared component the question concerns and would bias the comparison toward finding the three distinct. We report the mutually adjusted version in Table E3. We compared the three sets of estimates by the mean of their three pairwise Spearman correlations, and calibrated that value against a null in which they share one underlying pattern, taking the average of the three estimates for each protein as the common value, adding independent normal error with each condition’s own standard error, and recomputing the mean correlation over 10,000 simulations. The three conditions differed in direction for 13 of the 35 panel proteins. We did not adjust the primary models for medications, because metformin and statins mark treated disease and adjusting for them removes part of the exposure.

### E11. Tissue specificity

We took tissue specificity from the Human Protein Atlas version 25.1, using the RNA tissue specificity class and the single cell type expression values, and matched assays by gene symbol. The atlas calls a protein tissue enriched when its messenger RNA is at least four times higher in one tissue than in any other, group enriched when that holds for two to five tissues together, and tissue enhanced when one tissue exceeds the average of the rest by four times. Proteins meeting none of these have low tissue specificity. Cell type was assigned by the single cell cluster in which a protein was most highly expressed, the predominant cluster, rather than by any assignment to a cluster. We placed panel proteins within the distribution of estimates for assays of the same specificity class and repeated the compartment analysis after centering every estimate on its class mean, to separate compartment from tissue restriction.

### E12. Departure from linearity

We tested departure from linearity for every assay by an F test comparing the spline model with an otherwise identical model linear in BMI, and present the fitted relationships rather than summarizing them by a shape. Because non-linearity is easier to detect in a larger association, we matched each panel protein to platform assays whose absolute estimate fell within 0.02 SD of its own, took the proportion of those departing from linearity as the probability of detecting a departure in an association of that size, and obtained the expected number of non-linear panel proteins as the sum of those probabilities. We also read slopes over three segments of the BMI range bounded by the fifth, 30th, 70th and 95th percentiles and classified each departure as a reversal, steepening, flattening or a departure without a change of form, using thresholds fixed in advance. That classification is given in Table E5.

### E13. Genetically inferred ancestry

We tested whether the association differed across ancestry groups with a single model containing the BMI spline, indicators for the six groups, their fifteen interactions and the covariates above, testing the interaction terms jointly by F test, and derived each group’s contrast from that model. European ancestry is the reference because it is the largest group. Nine of the 18 proteins with heterogeneity exceeded a Bonferroni threshold of 0.0014, PAI-1, E-selectin, IL-8, CC16, IL-6, angiopoietin-2, ICAM-1, IL-10 and Gas6. Among the epithelial proteins, sRAGE and SP-A2 did not differ across groups (P = 0.29 and 0.93), CC16 did (P < 0.001) and was higher in participants of African ancestry and lower in the other five, and SP-D differed more weakly (P = 0.02). The alternative analysis, fitting each group separately and comparing estimates by Cochran’s Q, is given in Table E6. Because the 35 tests are correlated through shared biology and a shared exposure, a Bonferroni threshold is conservative, and we report it alongside the uncorrected P values rather than as the sole criterion.

### E14. Sensitivity analyses

Sensitivity analyses examined the addition of creatinine, the substitution of waist circumference for BMI, the removal of all thirteen conditions, the exclusion of participants admitted to hospital within 30 days of collection, the substitution of waist to hip ratio, restriction to participants with at least one diagnosis record, adjustment for statin and metformin exposure in the year before collection, identified from drug concepts for the active ingredient and every product containing it, and exclusion of participants the program flags as related to another participant at a kinship coefficient above 0.1. We evaluated waist circumference and waist to hip ratio between the values at the 34th and 86th percentiles of their own distributions, which covers the same part of the population as the BMI contrast. Results are given in Table E7.

**Supplementary tables**

Table E1. Comparison of assayed and unassayed participants within the long-read sequencing cohort, with models for selection.

| **Field** | **Value** |
| --- | --- |
| **A. Long-read frame, assayed against the rest** | |
| Participants, Not assayed | 5278 |
| Participants, Assayed | 9243 |
| Age, median years, Not assayed | 55 |
| Age, median years, Assayed | 56 |
| BMI, median kg/m2, Not assayed | 30.2 |
| BMI, median kg/m2, Assayed | 27.1 |
| BMI missing, percent, Not assayed | 0.7 |
| BMI missing, percent, Assayed | 0.6 |
| With any diagnosis record, percent, Not assayed | 81 |
| With any diagnosis record, percent, Assayed | 77.9 |
| **B. Long-read participants by genome center** | |
| Center BI, participants | 9922 |
| Center BI, percent assayed | 93 |
| All other centers combined, participants | 4599 |
| All other centers combined, percent assayed | fewer than 20 assayed |
| **C. Logistic model for being assayed, long read frame** | |
| Intercept, log odds | 1.9980 |
| Intercept, SE | 0.1019 |
| Intercept, z | 19.60 |
| Intercept, P | 1.56e-85 |
| Age, years, log odds | 0.0033 |
| Age, years, SE | 0.0011 |
| Age, years, z | 3.03 |
| Age, years, P | 0.00242 |
| BMI, kg/m2, log odds | -0.0609 |
| BMI, kg/m2, SE | 0.0026 |
| BMI, kg/m2, z | -23.58 |
| BMI, kg/m2, P | 6.25e-123 |
| BMI missing, log odds | -0.3722 |
| BMI missing, SE | 0.2143 |
| BMI missing, z | -1.74 |
| BMI missing, P | 0.0824 |
| Any diagnosis record, log odds | -0.0921 |
| Any diagnosis record, SE | 0.0453 |
| Any diagnosis record, z | -2.03 |
| Any diagnosis record, P | 0.0421 |
| Sex at birth | adjusted as a factor, coefficients not reported |
| c statistic | 0.667 |
| **D. Logistic model for being assayed, the center supplying the cohort** | |
| Intercept, log odds | 2.6989 |
| Intercept, SE | 0.2219 |
| Intercept, z | 12.16 |
| Intercept, P | 4.95e-34 |
| Age, years, log odds | -0.0072 |
| Age, years, SE | 0.0022 |
| Age, years, z | -3.18 |
| Age, years, P | 0.00148 |
| BMI, kg/m2, log odds | -0.0001 |
| BMI, kg/m2, SE | 0.0061 |
| BMI, kg/m2, z | -0.01 |
| BMI, kg/m2, P | 0.989 |
| BMI missing, log odds | -0.4834 |
| BMI missing, SE | 0.4057 |
| BMI missing, z | -1.19 |
| BMI missing, P | 0.233 |
| Any diagnosis record, log odds | 0.2938 |
| Any diagnosis record, SE | 0.0910 |
| Any diagnosis record, z | 3.23 |
| Any diagnosis record, P | 0.00125 |
| Sex at birth | adjusted as a factor, coefficients not reported |
| c statistic | 0.553 |
| **E. Proportion assayed by tenth of BMI, long read frame** | |
| Tenth 1, participants | 1451 |
| Tenth 1, percent assayed | 75.1 |
| Tenth 2, participants | 1460 |
| Tenth 2, percent assayed | 76.5 |
| Tenth 3, participants | 1455 |
| Tenth 3, percent assayed | 73.1 |
| Tenth 4, participants | 1405 |
| Tenth 4, percent assayed | 71.1 |
| Tenth 5, participants | 1443 |
| Tenth 5, percent assayed | 68.5 |
| Tenth 6, participants | 1443 |
| Tenth 6, percent assayed | 63.8 |
| Tenth 7, participants | 1445 |
| Tenth 7, percent assayed | 59.3 |
| Tenth 8, participants | 1440 |
| Tenth 8, percent assayed | 56.2 |
| Tenth 9, participants | 1450 |
| Tenth 9, percent assayed | 51.4 |
| Tenth 10, participants | 1436 |
| Tenth 10, percent assayed | 41.7 |
| **F. Proportion assayed by tenth of BMI, the center supplying the cohort** | |
| Tenth 1, participants | 987 |
| Tenth 1, percent assayed | 91.8 |
| Tenth 2, participants | 986 |
| Tenth 2, percent assayed | 92.7 |
| Tenth 3, participants | 986 |
| Tenth 3, percent assayed | 92.2 |
| Tenth 4, participants | 986 |
| Tenth 4, percent assayed | 93.7 |
| Tenth 5, participants | 986 |
| Tenth 5, percent assayed | 93.7 |
| Tenth 6, participants | 986 |
| Tenth 6, percent assayed | 94.3 |
| Tenth 7, participants | 986 |
| Tenth 7, percent assayed | 92.3 |
| Tenth 8, participants | 992 |
| Tenth 8, percent assayed | 93.9 |
| Tenth 9, participants | 982 |
| Tenth 9, percent assayed | 93.7 |
| Tenth 10, participants | 984 |
| Tenth 10, percent assayed | 92.2 |

*The long-read frame is the manifest of participants with long-read sequencing. Centers with fewer than 20 assayed participants are combined, and the combined row reports its size without a percentage, since a percentage of a known total is a count. Sex at birth was adjusted for in both models; its coefficients are not reported because the rare categories are small groups.*

Table E2. Concept identifiers for every variable drawn from the health record.

| **Variable** | **Concept ID** | **Concept name** | **Domain** | **Expansion** | **Rule** |
| --- | --- | --- | --- | --- | --- |
| Type 2 diabetes | 201826 | Type 2 diabetes mellitus | Condition occurrence | Concept and all descendants | Two occurrences on distinct dates before collection |
| Hypertension | 316866 | Hypertensive disorder | Condition occurrence | Concept and all descendants | Two occurrences on distinct dates before collection |
| Dyslipidemia | 432867 | Hyperlipidemia | Condition occurrence | Concept and all descendants | Two occurrences on distinct dates before collection |
| Heart failure | 316139 | Heart failure | Condition occurrence | Concept and all descendants | Two occurrences on distinct dates before collection |
| Atrial fibrillation | 313217 | Atrial fibrillation | Condition occurrence | Concept and all descendants | Two occurrences on distinct dates before collection |
| Chronic obstructive pulmonary disease | 255573 | Chronic obstructive pulmonary disease | Condition occurrence | Concept and all descendants | Two occurrences on distinct dates before collection |
| Interstitial lung disease | 4119786 | Interstitial lung disease | Condition occurrence | Concept and all descendants | Two occurrences on distinct dates before collection |
| Asthma | 317009 | Asthma | Condition occurrence | Concept and all descendants | Two occurrences on distinct dates before collection |
| Cancer | 443392 | Malignant neoplastic disease | Condition occurrence | Concept and all descendants | Two occurrences on distinct dates before collection |
| Obstructive sleep apnea | 442588 | Obstructive sleep apnea syndrome | Condition occurrence | Concept and all descendants | Two occurrences on distinct dates before collection |
| Disorder caused by alcohol | 36714559 | Disorder caused by alcohol | Condition occurrence | Concept and all descendants | Two occurrences on distinct dates before collection |
| Coronary artery disease | 317576 | Coronary arteriosclerosis | Condition occurrence | Concept and all descendants | Two occurrences on distinct dates before collection |
| Cerebrovascular disease | 381591 | Cerebrovascular disease | Condition occurrence | Concept and all descendants | Two occurrences on distinct dates before collection |
| Smoking, ever, question | 1585857 | Smoking: 100 Cigs Lifetime | Observation, survey | Source concept, no descendants | Response nearest enrollment |
| Smoking, ever, answer yes | 1585858 | 100 Cigs Lifetime: Yes | Observation, survey | Source concept, no descendants | Response nearest enrollment |
| Smoking, ever, answer no | 1585859 | 100 Cigs Lifetime: No | Observation, survey | Source concept, no descendants | Response nearest enrollment |
| Smoking, current, question | 1585860 | Smoking: Smoke Frequency | Observation, survey | Source concept, no descendants | Response nearest enrollment |
| Smoking, current, answer every day | 1585861 | Smoke Frequency: Every Day | Observation, survey | Source concept, no descendants | Response nearest enrollment |
| Smoking, current, answer some days | 1585862 | Smoke Frequency: Some Days | Observation, survey | Source concept, no descendants | Response nearest enrollment |
| Smoking, current, answer not at all | 1585863 | Smoke Frequency: Not At All | Observation, survey | Source concept, no descendants | Response nearest enrollment |
| Body mass index | 3038553 | Body mass index (BMI) [Ratio] | Measurement | Concept only | Physical examination, measurement type 44818701, value nearest collection |
| Waist circumference | 903135 | Computed waist circumference, mean of closest two measures | Measurement | Concept only | Physical examination, measurement type 44818701, value nearest collection |
| Hip circumference | 903136 | Computed hip circumference, mean of closest two measures | Measurement | Concept only | Physical examination, measurement type 44818701, value nearest collection |
| Systolic blood pressure | 903118 | Computed systolic blood pressure, mean of 2nd and 3rd measures | Measurement | Concept only | Physical examination, measurement type 44818701, value nearest collection |
| Diastolic blood pressure | 903115 | Computed diastolic blood pressure, mean of 2nd and 3rd measures | Measurement | Concept only | Physical examination, measurement type 44818701, value nearest collection |
| Serum creatinine | 3016723 | Creatinine [Mass/volume] in Serum or Plasma | Measurement | Concept only | Nearest value before collection, within two years |
| Inpatient visit | 9201 | Inpatient Visit | Visit occurrence | Concept only | Encounter spanning collection excluded; encounter within 30 days flagged |
| Emergency department and inpatient visit | 262 | Emergency Room and Inpatient Visit | Visit occurrence | Concept only | Encounter spanning collection excluded; encounter within 30 days flagged |
| Inpatient hospital | 8717 | Inpatient Hospital | Visit occurrence | Concept only | Encounter spanning collection excluded; encounter within 30 days flagged |
| Inpatient rehabilitation facility | 8920 | Comprehensive Inpatient Rehabilitation Facility | Visit occurrence | Concept only | Encounter spanning collection excluded; encounter within 30 days flagged |
| Statin | 1539403 | simvastatin | Drug exposure | Ingredient and all descendants | Any exposure in the year before collection, sensitivity analysis only |
| Statin | 1545958 | atorvastatin | Drug exposure | Ingredient and all descendants | Any exposure in the year before collection, sensitivity analysis only |
| Statin | 1510813 | rosuvastatin | Drug exposure | Ingredient and all descendants | Any exposure in the year before collection, sensitivity analysis only |
| Statin | 1551860 | pravastatin | Drug exposure | Ingredient and all descendants | Any exposure in the year before collection, sensitivity analysis only |
| Statin | 1592085 | lovastatin | Drug exposure | Ingredient and all descendants | Any exposure in the year before collection, sensitivity analysis only |
| Statin | 1549686 | fluvastatin | Drug exposure | Ingredient and all descendants | Any exposure in the year before collection, sensitivity analysis only |
| Statin | 40165636 | pitavastatin | Drug exposure | Ingredient and all descendants | Any exposure in the year before collection, sensitivity analysis only |
| Metformin | 1503297 | metformin | Drug exposure | Ingredient and all descendants | Any exposure in the year before collection, sensitivity analysis only |

*Every concept listed is a standard concept in the Observational Medical Outcomes Partnership vocabulary. Conditions are named by the concept queried.*

Table E3. Metabolic condition estimates under mutual adjustment.

| **Protein** | **Gene** | **Set** | **BMI 35 vs 25** | **Type 2 diabetes** | **Hypertension** | **Dyslipidemia** |
| --- | --- | --- | --- | --- | --- | --- |
| CC16 | SCGB1A1 | Epithelial | -0.081 | 0.120 (0.033) | 0.096 (0.027) | 0.001 (0.028) |
| SP-A2 | SFTPA2 | Epithelial | -0.084 | -0.055 (0.035) | -0.048 (0.030) | -0.014 (0.031) |
| SP-D | SFTPD | Epithelial | -0.083 | -0.009 (0.035) | 0.042 (0.029) | 0.016 (0.030) |
| sRAGE | AGER | Epithelial | -0.350 | 0.057 (0.033) | -0.001 (0.028) | -0.030 (0.029) |
| Gas6 | GAS6 | Endothelial activation and coagulation | 0.133 | 0.108 (0.035) | 0.099 (0.030) | -0.089 (0.030) |
| PAI-1 | SERPINE1 | Endothelial activation and coagulation | 0.292 | -0.007 (0.036) | 0.057 (0.030) | 0.068 (0.031) |
| protein C | PROC | Endothelial activation and coagulation | 0.176 | 0.057 (0.036) | -0.013 (0.030) | 0.228 (0.031) |
| thrombomodulin | THBD | Endothelial activation and coagulation | 0.041 | 0.191 (0.034) | 0.174 (0.029) | -0.053 (0.029) |
| vWF | VWF | Endothelial activation and coagulation | 0.215 | 0.040 (0.035) | 0.072 (0.030) | -0.020 (0.030) |
| ACE | ACE | Endothelial permeability | 0.025 | 0.090 (0.036) | 0.187 (0.030) | -0.054 (0.031) |
| RIPK3 | RIPK3 | Endothelial permeability | 0.164 | -0.006 (0.037) | -0.002 (0.031) | 0.044 (0.032) |
| TIMP-3 | TIMP3 | Endothelial permeability | 0.052 | 0.086 (0.036) | 0.022 (0.030) | -0.034 (0.031) |
| angiopoietin-1 | ANGPT1 | Endothelial permeability | -0.056 | 0.038 (0.036) | 0.010 (0.031) | 0.038 (0.032) |
| angiopoietin-2 | ANGPT2 | Endothelial permeability | 0.142 | 0.220 (0.035) | 0.071 (0.029) | -0.097 (0.030) |
| azurocidin | AZU1 | Endothelial permeability | 0.326 | 0.049 (0.036) | 0.074 (0.031) | 0.037 (0.031) |
| E-selectin | SELE | Inflammation | 0.465 | 0.255 (0.034) | 0.083 (0.029) | 0.010 (0.029) |
| G-CSF | CSF3 | Inflammation | 0.168 | 0.005 (0.034) | -0.021 (0.029) | 0.001 (0.030) |
| ICAM-1 | ICAM1 | Inflammation | 0.285 | 0.180 (0.034) | 0.050 (0.029) | -0.092 (0.030) |
| IL-10 | IL10 | Inflammation | 0.005 | 0.146 (0.036) | 0.052 (0.031) | -0.064 (0.031) |
| IL-17A | IL17A | Inflammation | 0.044 | 0.195 (0.037) | 0.041 (0.031) | -0.027 (0.032) |
| IL-18 | IL18 | Inflammation | 0.250 | 0.131 (0.036) | 0.101 (0.030) | -0.018 (0.031) |
| IL-1B | IL1B | Inflammation | 0.189 | 0.030 (0.037) | 0.012 (0.031) | -0.026 (0.032) |
| IL-6 | IL6 | Inflammation | 0.534 | 0.118 (0.033) | 0.083 (0.028) | -0.075 (0.029) |
| IL-8 | CXCL8 | Inflammation | -0.047 | 0.281 (0.034) | 0.047 (0.029) | -0.046 (0.030) |
| MPO | MPO | Inflammation | 0.374 | 0.121 (0.036) | 0.089 (0.030) | 0.042 (0.031) |
| NAMPT | NAMPT | Inflammation | 0.309 | 0.041 (0.037) | 0.059 (0.031) | 0.039 (0.032) |
| TNF | TNF | Inflammation | 0.072 | 0.176 (0.036) | 0.074 (0.031) | 0.001 (0.031) |
| elafin | PI3 | Inflammation | 0.175 | 0.141 (0.033) | 0.119 (0.028) | -0.059 (0.029) |
| elastase | ELANE | Inflammation | 0.270 | 0.066 (0.036) | 0.091 (0.031) | -0.001 (0.031) |
| sTNFR-1 | TNFRSF1A | Inflammation | 0.326 | 0.313 (0.032) | 0.216 (0.027) | -0.115 (0.028) |
| sTNFR-2 | TNFRSF1B | Inflammation | 0.286 | 0.309 (0.033) | 0.169 (0.028) | -0.095 (0.028) |
| suPAR | PLAUR | Inflammation | 0.225 | 0.230 (0.033) | 0.194 (0.028) | -0.083 (0.028) |
| BNP | NPPB | Myocardial | -0.103 | 0.012 (0.031) | 0.112 (0.027) | -0.148 (0.027) |
| troponin I | TNNI3 | Myocardial | 0.067 | 0.008 (0.037) | 0.037 (0.031) | -0.041 (0.032) |
| troponin T | TNNT2 | Myocardial | -0.029 | -0.014 (0.037) | -0.000 (0.031) | 0.059 (0.032) |

*Values are the estimate and its standard error in standard deviation units, with body mass index held fixed by the same spline.*

Table E4. Body size associations for the 35 panel proteins, with the percentile of each estimate within the platform distribution.

| **Protein** | **Gene** | **Set** | **Estimate** | **CI** | **Percentile** | **n** |
| --- | --- | --- | --- | --- | --- | --- |
| sRAGE | AGER | Epithelial | -0.350 | -0.404 to -0.297 | 1.2 | 8673 |
| BNP | NPPB | Myocardial | -0.103 | -0.153 to -0.053 | 6.2 | 8607 |
| SP-A2 | SFTPA2 | Epithelial | -0.084 | -0.140 to -0.027 | 7.1 | 8607 |
| SP-D | SFTPD | Epithelial | -0.083 | -0.138 to -0.027 | 7.1 | 8673 |
| CC16 | SCGB1A1 | Epithelial | -0.081 | -0.133 to -0.029 | 7.2 | 8673 |
| angiopoietin-1 | ANGPT1 | Endothelial permeability | -0.056 | -0.115 to 0.002 | 8.8 | 8673 |
| IL-8 | CXCL8 | Inflammation | -0.047 | -0.103 to 0.008 | 9.8 | 8613 |
| troponin T | TNNT2 | Myocardial | -0.029 | -0.088 to 0.030 | 12.4 | 8599 |
| IL-10 | IL10 | Inflammation | 0.005 | -0.053 to 0.064 | 22.7 | 8607 |
| ACE | ACE | Endothelial permeability | 0.025 | -0.033 to 0.082 | 32.1 | 8665 |
| thrombomodulin | THBD | Endothelial activation and coagulation | 0.041 | -0.014 to 0.096 | 39.8 | 8673 |
| IL-17A | IL17A | Inflammation | 0.044 | -0.014 to 0.103 | 41.2 | 8607 |
| TIMP-3 | TIMP3 | Endothelial permeability | 0.052 | -0.006 to 0.109 | 44.9 | 8673 |
| troponin I | TNNI3 | Myocardial | 0.067 | 0.008 to 0.125 | 50.9 | 8607 |
| TNF | TNF | Inflammation | 0.072 | 0.013 to 0.130 | 52.6 | 8607 |
| Gas6 | GAS6 | Endothelial activation and coagulation | 0.133 | 0.076 to 0.189 | 69.6 | 8673 |
| angiopoietin-2 | ANGPT2 | Endothelial permeability | 0.142 | 0.087 to 0.198 | 71.4 | 8613 |
| RIPK3 | RIPK3 | Endothelial permeability | 0.164 | 0.105 to 0.222 | 75.8 | 8599 |
| G-CSF | CSF3 | Inflammation | 0.168 | 0.113 to 0.223 | 76.6 | 8607 |
| elafin | PI3 | Inflammation | 0.175 | 0.122 to 0.228 | 78.0 | 8673 |
| protein C | PROC | Endothelial activation and coagulation | 0.176 | 0.119 to 0.233 | 78.2 | 8665 |
| IL-1B | IL1B | Inflammation | 0.189 | 0.130 to 0.248 | 80.8 | 8607 |
| vWF | VWF | Endothelial activation and coagulation | 0.215 | 0.159 to 0.272 | 85.5 | 8673 |
| suPAR | PLAUR | Inflammation | 0.225 | 0.172 to 0.277 | 87.1 | 8673 |
| IL-18 | IL18 | Inflammation | 0.250 | 0.193 to 0.308 | 90.5 | 8673 |
| elastase | ELANE | Inflammation | 0.270 | 0.212 to 0.328 | 92.5 | 8665 |
| ICAM-1 | ICAM1 | Inflammation | 0.285 | 0.231 to 0.340 | 93.9 | 8665 |
| sTNFR-2 | TNFRSF1B | Inflammation | 0.286 | 0.233 to 0.338 | 94.0 | 8673 |
| PAI-1 | SERPINE1 | Endothelial activation and coagulation | 0.292 | 0.234 to 0.349 | 94.5 | 8673 |
| NAMPT | NAMPT | Inflammation | 0.309 | 0.250 to 0.368 | 95.4 | 8607 |
| sTNFR-1 | TNFRSF1A | Inflammation | 0.326 | 0.274 to 0.377 | 96.3 | 8673 |
| azurocidin | AZU1 | Endothelial permeability | 0.326 | 0.268 to 0.384 | 96.3 | 8673 |
| MPO | MPO | Inflammation | 0.374 | 0.317 to 0.431 | 98.0 | 8673 |
| E-selectin | SELE | Inflammation | 0.465 | 0.411 to 0.520 | 99.2 | 8673 |
| IL-6 | IL6 | Inflammation | 0.534 | 0.481 to 0.588 | 99.5 | 8613 |

*Estimates are the difference in standardized concentration between a body mass index of 35 and 25.*

Table E5. Classification of departures from linearity by shape.

| **Protein** | **Gene** | **Set** | **BMI 35 vs 25** | **Slope, lower** | **Slope, middle** | **Slope, upper** | **P for nonlinearity** | **Form** | **n** |
| --- | --- | --- | --- | --- | --- | --- | --- | --- | --- |
| angiopoietin-2 | ANGPT2 | Endothelial permeability | 0.142 | -0.0378 | 0.0065 | 0.0243 | 2.6e-21 | reversal | 8613 |
| sRAGE | AGER | Epithelial | -0.350 | -0.0598 | -0.0423 | -0.0204 | 3.1e-17 | gentle | 8673 |
| Gas6 | GAS6 | Endothelial activation and coagulation | 0.133 | -0.0402 | 0.0069 | 0.0193 | 1.7e-16 | reversal | 8673 |
| protein C | PROC | Endothelial activation and coagulation | 0.176 | 0.0404 | 0.0239 | 0.0051 | 7.5e-12 | gentle | 8665 |
| suPAR | PLAUR | Inflammation | 0.225 | -0.0161 | 0.0181 | 0.0261 | 1.4e-09 | reversal | 8673 |
| MPO | MPO | Inflammation | 0.374 | 0.0154 | 0.0415 | 0.0214 | 1.6e-09 | gentle | 8673 |
| azurocidin | AZU1 | Endothelial permeability | 0.326 | 0.0092 | 0.0366 | 0.0165 | 2.1e-09 | flattening | 8673 |
| BNP | NPPB | Myocardial | -0.103 | -0.0294 | -0.0152 | -0.0009 | 2.4e-09 | gentle | 8607 |
| NAMPT | NAMPT | Inflammation | 0.309 | 0.0075 | 0.0349 | 0.0149 | 4e-09 | flattening | 8607 |
| E-selectin | SELE | Inflammation | 0.465 | 0.0258 | 0.0501 | 0.0324 | 2e-08 | gentle | 8673 |
| IL-8 | CXCL8 | Inflammation | -0.047 | -0.0285 | -0.0094 | 0.0031 | 1.4e-07 | steepening | 8613 |
| IL-6 | IL6 | Inflammation | 0.534 | 0.0189 | 0.0543 | 0.0434 | 9.6e-07 | gentle | 8613 |
| sTNFR-2 | TNFRSF1B | Inflammation | 0.286 | -0.0042 | 0.0264 | 0.0276 | 2.8e-05 | gentle | 8673 |
| elafin | PI3 | Inflammation | 0.175 | -0.0130 | 0.0153 | 0.0168 | 0.00012 | reversal | 8673 |
| ACE | ACE | Endothelial permeability | 0.025 | -0.0263 | -0.0005 | 0.0043 | 0.00016 | steepening | 8665 |
| RIPK3 | RIPK3 | Endothelial permeability | 0.164 | 0.0060 | 0.0194 | 0.0059 | 0.00021 | flattening | 8599 |
| SP-D | SFTPD | Epithelial | -0.083 | -0.0325 | -0.0112 | -0.0055 | 0.00037 | steepening | 8673 |
| ICAM-1 | ICAM1 | Inflammation | 0.285 | 0.0015 | 0.0260 | 0.0296 | 4e-04 | gentle | 8665 |
| elastase | ELANE | Inflammation | 0.270 | 0.0151 | 0.0293 | 0.0181 | 0.0022 | gentle | 8665 |
| sTNFR-1 | TNFRSF1A | Inflammation | 0.326 | 0.0069 | 0.0317 | 0.0294 | 0.0023 | gentle | 8673 |
| PAI-1 | SERPINE1 | Endothelial activation and coagulation | 0.292 | 0.0205 | 0.0316 | 0.0207 | 0.0034 | gentle | 8673 |
| vWF | VWF | Endothelial activation and coagulation | 0.215 | -0.0026 | 0.0201 | 0.0201 | 0.0098 | gentle | 8673 |
| IL-17A | IL17A | Inflammation | 0.044 | -0.0096 | 0.0018 | 0.0085 | 0.012 | reversal | 8607 |
| IL-18 | IL18 | Inflammation | 0.250 | 0.0082 | 0.0260 | 0.0187 | 0.023 | gentle | 8673 |
| CC16 | SCGB1A1 | Epithelial | -0.081 | -0.0187 | -0.0101 | -0.0048 | 0.034 | gentle | 8673 |
| IL-1B | IL1B | Inflammation | 0.189 | -0.0001 | 0.0188 | 0.0149 | 0.06 | linear | 8607 |
| IL-10 | IL10 | Inflammation | 0.005 | -0.0117 | -0.0013 | 0.0030 | 0.084 | linear | 8607 |
| troponin T | TNNT2 | Myocardial | -0.029 | 0.0134 | -0.0028 | 0.0002 | 0.14 | linear | 8599 |
| TNF | TNF | Inflammation | 0.072 | -0.0071 | 0.0059 | 0.0074 | 0.16 | linear | 8607 |
| thrombomodulin | THBD | Endothelial activation and coagulation | 0.041 | -0.0016 | 0.0027 | 0.0068 | 0.24 | linear | 8673 |
| TIMP-3 | TIMP3 | Endothelial permeability | 0.052 | -0.0024 | 0.0038 | 0.0074 | 0.27 | linear | 8673 |
| angiopoietin-1 | ANGPT1 | Endothelial permeability | -0.056 | 0.0009 | -0.0065 | -0.0019 | 0.34 | linear | 8673 |
| SP-A2 | SFTPA2 | Epithelial | -0.084 | -0.0036 | -0.0083 | -0.0074 | 0.83 | linear | 8607 |
| troponin I | TNNI3 | Myocardial | 0.067 | 0.0076 | 0.0072 | 0.0054 | 0.83 | linear | 8607 |
| G-CSF | CSF3 | Inflammation | 0.168 | 0.0161 | 0.0165 | 0.0175 | 0.94 | linear | 8607 |

*Slopes are per unit of body mass index over thirds of its range bounded by the fifth, 30th, 70th and 95th percentiles.*

Table E6. Ancestry specific estimates fitted separately, with Cochran’s Q.

| **Protein** | **Gene** | **Set** | **BMI 35 vs 25** | **afr** | **amr** | **eas** | **eur** | **mid** | **sas** | **k** | **Q** | **df** | **P** | **I²** |
| --- | --- | --- | --- | --- | --- | --- | --- | --- | --- | --- | --- | --- | --- | --- |
| PAI-1 | SERPINE1 | Endothelial activation and coagulation | 0.292 | 0.027 (0.064) | 0.314 (0.058) | 0.463 (0.076) | 0.403 (0.052) | 0.170 (0.136) | 0.424 (0.083) | 6 | 29.8 | 5 | 1.6e-05 | 83 |
| IL-8 | CXCL8 | Inflammation | -0.047 | -0.278 (0.060) | -0.077 (0.056) | 0.134 (0.074) | 0.069 (0.052) | 0.169 (0.131) | 0.038 (0.080) | 6 | 29.1 | 5 | 2.2e-05 | 83 |
| IL-10 | IL10 | Inflammation | 0.005 | -0.226 (0.064) | -0.026 (0.060) | 0.173 (0.077) | 0.073 (0.054) | 0.020 (0.139) | 0.264 (0.084) | 6 | 28.5 | 5 | 2.8e-05 | 82 |
| ICAM-1 | ICAM1 | Inflammation | 0.285 | 0.129 (0.063) | 0.316 (0.057) | 0.385 (0.075) | 0.263 (0.051) | 0.545 (0.130) | 0.558 (0.080) | 6 | 23 | 5 | 0.00034 | 78 |
| TIMP-3 | TIMP3 | Endothelial permeability | 0.052 | -0.095 (0.065) | 0.016 (0.059) | 0.318 (0.077) | 0.084 (0.054) | -0.016 (0.137) | 0.206 (0.085) | 6 | 20.8 | 5 | 9e-04 | 76 |
| E-selectin | SELE | Inflammation | 0.465 | 0.276 (0.063) | 0.425 (0.056) | 0.660 (0.072) | 0.533 (0.051) | 0.614 (0.128) | 0.525 (0.080) | 6 | 20.4 | 5 | 0.0011 | 75 |
| IL-6 | IL6 | Inflammation | 0.534 | 0.367 (0.061) | 0.591 (0.055) | 0.605 (0.073) | 0.522 (0.049) | 0.752 (0.125) | 0.737 (0.076) | 6 | 19.1 | 5 | 0.0018 | 74 |
| CC16 | SCGB1A1 | Epithelial | -0.081 | 0.093 (0.059) | -0.085 (0.055) | -0.236 (0.072) | -0.105 (0.047) | -0.268 (0.127) | -0.194 (0.079) | 6 | 17.4 | 5 | 0.0039 | 71 |
| suPAR | PLAUR | Inflammation | 0.225 | 0.102 (0.061) | 0.216 (0.054) | 0.313 (0.075) | 0.254 (0.046) | 0.163 (0.129) | 0.444 (0.076) | 6 | 14 | 5 | 0.016 | 64 |
| angiopoietin-1 | ANGPT1 | Endothelial permeability | -0.056 | -0.178 (0.064) | -0.025 (0.059) | 0.151 (0.078) | -0.013 (0.054) | -0.192 (0.134) | 0.034 (0.085) | 6 | 12.9 | 5 | 0.024 | 61 |
| Gas6 | GAS6 | Endothelial activation and coagulation | 0.133 | 0.001 (0.064) | 0.203 (0.058) | 0.168 (0.076) | 0.098 (0.052) | 0.111 (0.133) | 0.341 (0.082) | 6 | 12.8 | 5 | 0.026 | 61 |
| elastase | ELANE | Inflammation | 0.270 | 0.248 (0.065) | 0.146 (0.060) | 0.431 (0.077) | 0.228 (0.053) | 0.337 (0.138) | 0.386 (0.083) | 6 | 11.6 | 5 | 0.041 | 57 |
| angiopoietin-2 | ANGPT2 | Endothelial permeability | 0.142 | 0.060 (0.063) | 0.028 (0.058) | 0.193 (0.075) | 0.211 (0.050) | 0.305 (0.127) | 0.236 (0.083) | 6 | 10.8 | 5 | 0.055 | 54 |
| SP-D | SFTPD | Epithelial | -0.083 | -0.197 (0.063) | -0.024 (0.058) | 0.062 (0.077) | -0.033 (0.052) | -0.257 (0.135) | -0.133 (0.083) | 6 | 10.6 | 5 | 0.059 | 53 |
| sTNFR-1 | TNFRSF1A | Inflammation | 0.326 | 0.229 (0.060) | 0.311 (0.054) | 0.389 (0.072) | 0.308 (0.046) | 0.341 (0.122) | 0.523 (0.076) | 6 | 10.3 | 5 | 0.067 | 52 |
| vWF | VWF | Endothelial activation and coagulation | 0.215 | 0.111 (0.064) | 0.150 (0.058) | 0.162 (0.076) | 0.276 (0.052) | 0.317 (0.128) | 0.364 (0.083) | 6 | 9.6 | 5 | 0.087 | 48 |
| sTNFR-2 | TNFRSF1B | Inflammation | 0.286 | 0.194 (0.061) | 0.249 (0.055) | 0.339 (0.072) | 0.292 (0.047) | 0.271 (0.124) | 0.480 (0.077) | 6 | 9.5 | 5 | 0.092 | 47 |
| IL-17A | IL17A | Inflammation | 0.044 | 0.031 (0.065) | 0.012 (0.060) | -0.031 (0.078) | 0.149 (0.053) | 0.123 (0.138) | -0.096 (0.085) | 6 | 8.2 | 5 | 0.14 | 39 |
| IL-18 | IL18 | Inflammation | 0.250 | 0.123 (0.064) | 0.266 (0.058) | 0.366 (0.076) | 0.247 (0.052) | 0.087 (0.130) | 0.309 (0.083) | 6 | 8.4 | 5 | 0.14 | 41 |
| thrombomodulin | THBD | Endothelial activation and coagulation | 0.041 | 0.029 (0.062) | 0.010 (0.054) | 0.142 (0.073) | -0.016 (0.050) | 0.050 (0.131) | 0.213 (0.079) | 6 | 8.3 | 5 | 0.14 | 39 |
| sRAGE | AGER | Epithelial | -0.350 | -0.251 (0.062) | -0.313 (0.058) | -0.355 (0.076) | -0.412 (0.051) | -0.421 (0.132) | -0.505 (0.082) | 6 | 8.1 | 5 | 0.15 | 38 |
| RIPK3 | RIPK3 | Endothelial permeability | 0.164 | 0.086 (0.065) | 0.119 (0.060) | 0.281 (0.078) | 0.182 (0.054) | -0.004 (0.139) | 0.288 (0.085) | 6 | 7.9 | 5 | 0.16 | 37 |
| NAMPT | NAMPT | Inflammation | 0.309 | 0.228 (0.065) | 0.291 (0.059) | 0.348 (0.078) | 0.269 (0.053) | 0.265 (0.138) | 0.501 (0.084) | 6 | 7.7 | 5 | 0.17 | 35 |
| ACE | ACE | Endothelial permeability | 0.025 | 0.023 (0.064) | 0.116 (0.057) | -0.026 (0.078) | -0.064 (0.053) | 0.085 (0.134) | 0.014 (0.082) | 6 | 5.9 | 5 | 0.31 | 16 |
| SP-A2 | SFTPA2 | Epithelial | -0.084 | -0.187 (0.062) | -0.109 (0.057) | -0.074 (0.078) | -0.033 (0.052) | -0.002 (0.136) | 0.028 (0.084) | 6 | 6 | 5 | 0.31 | 17 |
| BNP | NPPB | Myocardial | -0.103 | -0.163 (0.059) | -0.073 (0.054) | -0.086 (0.070) | -0.077 (0.044) | -0.175 (0.118) | -0.246 (0.072) | 6 | 5.7 | 5 | 0.33 | 13 |
| G-CSF | CSF3 | Inflammation | 0.168 | 0.186 (0.062) | 0.232 (0.057) | 0.049 (0.076) | 0.197 (0.053) | 0.330 (0.130) | 0.154 (0.082) | 6 | 5.3 | 5 | 0.38 | 6 |
| azurocidin | AZU1 | Endothelial permeability | 0.326 | 0.274 (0.065) | 0.279 (0.059) | 0.404 (0.077) | 0.282 (0.053) | 0.477 (0.136) | 0.408 (0.084) | 6 | 5.1 | 5 | 0.4 | 2 |
| elafin | PI3 | Inflammation | 0.175 | 0.055 (0.063) | 0.160 (0.057) | 0.249 (0.073) | 0.202 (0.049) | 0.158 (0.126) | 0.156 (0.080) | 6 | 5 | 5 | 0.42 | 0 |
| MPO | MPO | Inflammation | 0.374 | 0.381 (0.064) | 0.332 (0.059) | 0.387 (0.077) | 0.317 (0.052) | 0.457 (0.137) | 0.480 (0.083) | 6 | 3.6 | 5 | 0.6 | 0 |
| IL-1B | IL1B | Inflammation | 0.189 | 0.172 (0.065) | 0.209 (0.060) | 0.253 (0.078) | 0.157 (0.054) | 0.031 (0.139) | 0.260 (0.085) | 6 | 3.2 | 5 | 0.67 | 0 |
| troponin T | TNNT2 | Myocardial | -0.029 | 0.030 (0.065) | -0.030 (0.060) | 0.060 (0.079) | -0.060 (0.054) | -0.110 (0.138) | -0.080 (0.085) | 6 | 3.1 | 5 | 0.68 | 0 |
| protein C | PROC | Endothelial activation and coagulation | 0.176 | 0.244 (0.063) | 0.138 (0.059) | 0.168 (0.076) | 0.195 (0.052) | 0.170 (0.135) | 0.148 (0.082) | 6 | 1.8 | 5 | 0.88 | 0 |
| TNF | TNF | Inflammation | 0.072 | 0.107 (0.065) | 0.077 (0.059) | 0.091 (0.078) | 0.047 (0.054) | -0.034 (0.135) | 0.110 (0.084) | 6 | 1.4 | 5 | 0.93 | 0 |
| troponin I | TNNI3 | Myocardial | 0.067 | 0.046 (0.064) | 0.033 (0.060) | 0.064 (0.078) | 0.078 (0.054) | 0.117 (0.138) | 0.108 (0.085) | 6 | 0.8 | 5 | 0.98 | 0 |

*Values are the estimate and its standard error in standard deviation units. k, groups contributing; Q, Cochran’s Q; df, degrees of freedom; P, P value for heterogeneity; I², percentage of variation across groups beyond sampling error. afr, African; amr, Admixed American; eas, East Asian; eur, European; mid, Middle Eastern; sas, South Asian. Participants contributing to each column: afr 1,427 to 1,445; amr 1,839 to 1,858; eas 1,217 to 1,235; eur 2,566 to 2,590; mid 405 to 410; sas 1,131 to 1,140.*

Table E7. Sensitivity analyses, showing the estimate for every panel protein under each alternative specification.

| **Protein** | **Set** | **Primary** | **Creatinine** | **No conditions** | **No recent admission** | **Records only** | **Statin, metformin** | **BMI in subset** | **Waist** | **Waist to hip** | **Unrelated only** | **Plate** | **Batch** | **HC3 95% CI** |
| --- | --- | --- | --- | --- | --- | --- | --- | --- | --- | --- | --- | --- | --- | --- |
| CC16 | Epithelial | -0.081 | -0.089 | -0.045 | -0.075 | -0.088 | -0.078 | -0.076 | -0.028 | -0.082 | -0.070 | -0.081 | -0.081 | -0.134 to -0.028 |
| SP-A2 | Epithelial | -0.084 | -0.009 | -0.085 | -0.089 | -0.068 | -0.082 | -0.098 | -0.033 | 0.039 | -0.067 | -0.083 | -0.084 | -0.142 to -0.026 |
| SP-D | Epithelial | -0.083 | -0.106 | -0.077 | -0.083 | -0.091 | -0.085 | -0.093 | -0.069 | -0.007 | -0.084 | -0.088 | -0.083 | -0.139 to -0.027 |
| sRAGE | Epithelial | -0.350 | -0.360 | -0.331 | -0.350 | -0.370 | -0.346 | -0.351 | -0.288 | -0.197 | -0.349 | -0.352 | -0.350 | -0.405 to -0.296 |
| ACE | Endothelial permeability | 0.025 | -0.001 | 0.063 | 0.025 | 0.015 | 0.025 | 0.024 | -0.026 | 0.005 | 0.038 | 0.025 | 0.025 | -0.032 to 0.082 |
| RIPK3 | Endothelial permeability | 0.164 | 0.136 | 0.173 | 0.162 | 0.153 | 0.162 | 0.157 | 0.137 | 0.116 | 0.177 | 0.166 | 0.164 | 0.105 to 0.222 |
| TIMP-3 | Endothelial permeability | 0.052 | 0.055 | 0.050 | 0.055 | 0.048 | 0.052 | 0.046 | 0.068 | 0.077 | 0.053 | 0.053 | 0.051 | -0.005 to 0.109 |
| angiopoietin-1 | Endothelial permeability | -0.056 | -0.074 | -0.059 | -0.058 | -0.055 | -0.059 | -0.054 | -0.042 | -0.002 | -0.060 | -0.053 | -0.057 | -0.114 to 0.002 |
| angiopoietin-2 | Endothelial permeability | 0.142 | 0.163 | 0.176 | 0.143 | 0.124 | 0.143 | 0.206 | 0.239 | 0.193 | 0.140 | 0.144 | 0.142 | 0.087 to 0.198 |
| azurocidin | Endothelial permeability | 0.326 | 0.273 | 0.344 | 0.327 | 0.319 | 0.323 | 0.321 | 0.246 | 0.179 | 0.345 | 0.328 | 0.326 | 0.266 to 0.386 |
| Gas6 | Endothelial activation and coagulation | 0.133 | 0.108 | 0.149 | 0.131 | 0.139 | 0.136 | 0.125 | 0.092 | 0.095 | 0.130 | 0.130 | 0.133 | 0.077 to 0.189 |
| PAI-1 | Endothelial activation and coagulation | 0.292 | 0.272 | 0.295 | 0.293 | 0.293 | 0.289 | 0.283 | 0.273 | 0.244 | 0.297 | 0.301 | 0.291 | 0.235 to 0.348 |
| protein C | Endothelial activation and coagulation | 0.176 | 0.140 | 0.181 | 0.181 | 0.146 | 0.172 | 0.180 | 0.159 | 0.082 | 0.173 | 0.180 | 0.176 | 0.118 to 0.234 |
| thrombomodulin | Endothelial activation and coagulation | 0.041 | 0.008 | 0.084 | 0.042 | 0.026 | 0.045 | 0.039 | 0.067 | 0.065 | 0.044 | 0.037 | 0.041 | -0.012 to 0.094 |
| vWF | Endothelial activation and coagulation | 0.215 | 0.253 | 0.231 | 0.212 | 0.196 | 0.217 | 0.225 | 0.223 | 0.124 | 0.209 | 0.217 | 0.215 | 0.159 to 0.271 |
| E-selectin | Inflammation | 0.465 | 0.473 | 0.499 | 0.461 | 0.476 | 0.462 | 0.470 | 0.462 | 0.407 | 0.475 | 0.466 | 0.465 | 0.410 to 0.521 |
| G-CSF | Inflammation | 0.168 | 0.190 | 0.170 | 0.166 | 0.201 | 0.169 | 0.190 | 0.194 | 0.129 | 0.147 | 0.172 | 0.168 | 0.114 to 0.222 |
| ICAM-1 | Inflammation | 0.285 | 0.269 | 0.302 | 0.284 | 0.275 | 0.285 | 0.280 | 0.272 | 0.266 | 0.291 | 0.288 | 0.285 | 0.230 to 0.341 |
| IL-10 | Inflammation | 0.005 | 0.064 | 0.031 | 0.008 | 0.031 | 0.005 | -0.014 | -0.009 | 0.060 | 0.005 | 0.004 | 0.005 | -0.050 to 0.061 |
| IL-17A | Inflammation | 0.044 | 0.102 | 0.063 | 0.037 | 0.061 | 0.046 | 0.028 | 0.065 | 0.112 | 0.036 | 0.049 | 0.044 | -0.013 to 0.102 |
| IL-18 | Inflammation | 0.250 | 0.175 | 0.272 | 0.248 | 0.229 | 0.248 | 0.241 | 0.247 | 0.242 | 0.257 | 0.254 | 0.250 | 0.193 to 0.307 |
| IL-1B | Inflammation | 0.189 | 0.148 | 0.183 | 0.191 | 0.190 | 0.189 | 0.191 | 0.134 | 0.081 | 0.188 | 0.189 | 0.189 | 0.130 to 0.248 |
| IL-6 | Inflammation | 0.534 | 0.494 | 0.545 | 0.536 | 0.534 | 0.534 | 0.537 | 0.508 | 0.394 | 0.529 | 0.537 | 0.534 | 0.482 to 0.586 |
| IL-8 | Inflammation | -0.047 | -0.027 | -0.018 | -0.048 | -0.042 | -0.050 | -0.047 | -0.038 | 0.057 | -0.046 | -0.049 | -0.048 | -0.102 to 0.007 |
| MPO | Inflammation | 0.374 | 0.310 | 0.401 | 0.381 | 0.360 | 0.372 | 0.373 | 0.323 | 0.216 | 0.397 | 0.379 | 0.374 | 0.316 to 0.432 |
| NAMPT | Inflammation | 0.309 | 0.227 | 0.327 | 0.311 | 0.306 | 0.307 | 0.320 | 0.222 | 0.175 | 0.329 | 0.312 | 0.309 | 0.249 to 0.369 |
| TNF | Inflammation | 0.072 | 0.089 | 0.095 | 0.073 | 0.064 | 0.070 | 0.081 | 0.047 | 0.062 | 0.066 | 0.074 | 0.072 | 0.013 to 0.130 |
| elafin | Inflammation | 0.175 | 0.118 | 0.215 | 0.185 | 0.164 | 0.179 | 0.175 | 0.200 | 0.187 | 0.164 | 0.176 | 0.174 | 0.122 to 0.228 |
| elastase | Inflammation | 0.270 | 0.212 | 0.288 | 0.273 | 0.252 | 0.269 | 0.274 | 0.236 | 0.162 | 0.289 | 0.270 | 0.270 | 0.206 to 0.334 |
| sTNFR-1 | Inflammation | 0.326 | 0.249 | 0.387 | 0.332 | 0.315 | 0.329 | 0.319 | 0.357 | 0.278 | 0.314 | 0.325 | 0.325 | 0.275 to 0.377 |
| sTNFR-2 | Inflammation | 0.286 | 0.227 | 0.347 | 0.283 | 0.278 | 0.288 | 0.289 | 0.306 | 0.256 | 0.281 | 0.285 | 0.286 | 0.232 to 0.339 |
| suPAR | Inflammation | 0.225 | 0.179 | 0.269 | 0.225 | 0.209 | 0.226 | 0.227 | 0.248 | 0.271 | 0.219 | 0.224 | 0.224 | 0.175 to 0.274 |
| BNP | Myocardial | -0.103 | -0.142 | -0.085 | -0.096 | -0.116 | -0.099 | -0.107 | -0.074 | -0.062 | -0.112 | -0.107 | -0.103 | -0.156 to -0.050 |
| troponin I | Myocardial | 0.067 | 0.094 | 0.079 | 0.063 | 0.068 | 0.068 | 0.073 | 0.113 | 0.105 | 0.069 | 0.062 | 0.067 | 0.009 to 0.125 |
| troponin T | Myocardial | -0.029 | -0.035 | -0.028 | -0.029 | -0.020 | -0.030 | -0.039 | -0.027 | 0.031 | -0.040 | -0.027 | -0.029 | -0.087 to 0.028 |

*Every column is the same contrast, a body mass index of 35 against 25, in standard deviation units. Columns are, in order: the primary model; creatinine added; all thirteen chronic conditions removed; participants admitted to hospital within 30 days of collection excluded; restricted to participants with at least one diagnosis record; statin and metformin exposure in the year before collection adjusted for; body mass index refitted in the subset with a computable waist to hip ratio, with waist circumference and waist to hip ratio substituted as the exposure in that same subset; participants the program flags as related excluded; plate as a fixed effect; batch adjusted; and the 95% confidence interval from HC3 standard errors. Gene symbols are given in Table E4. Participants contributing to each column: Creatinine added 3,934 to 3,969; No chronic conditions 8,599 to 8,673; No admission within 30 days 8,349 to 8,424; Diagnosis records only 7,161 to 7,227; Statin and metformin adjusted 8,599 to 8,673; Waist subset 7,839; Unrelated only 7,709 to 7,774.*

Table E8. Estimates in the metabolically healthy stratum.

| Protein | Gene | Set | Primary | Metabolically healthy | With a diagnosis record |
| --- | --- | --- | --- | --- | --- |
| CC16 | SCGB1A1 | Epithelial | -0.081 | -0.125 | -0.152 |
| SP-A2 | SFTPA2 | Epithelial | -0.084 | -0.106 | -0.086 |
| SP-D | SFTPD | Epithelial | -0.083 | -0.111 | -0.136 |
| sRAGE | AGER | Epithelial | -0.350 | -0.367 | -0.417 |
| Gas6 | GAS6 | Endothelial activation and coagulation | 0.133 | 0.128 | 0.129 |
| PAI-1 | SERPINE1 | Endothelial activation and coagulation | 0.292 | 0.304 | 0.302 |
| protein C | PROC | Endothelial activation and coagulation | 0.176 | 0.233 | 0.201 |
| thrombomodulin | THBD | Endothelial activation and coagulation | 0.041 | 0.061 | 0.030 |
| vWF | VWF | Endothelial activation and coagulation | 0.215 | 0.181 | 0.131 |
| ACE | ACE | Endothelial permeability | 0.025 | 0.044 | 0.027 |
| RIPK3 | RIPK3 | Endothelial permeability | 0.164 | 0.196 | 0.180 |
| TIMP-3 | TIMP3 | Endothelial permeability | 0.052 | 0.066 | 0.056 |
| angiopoietin-1 | ANGPT1 | Endothelial permeability | -0.056 | -0.057 | -0.066 |
| angiopoietin-2 | ANGPT2 | Endothelial permeability | 0.142 | 0.096 | 0.058 |
| azurocidin | AZU1 | Endothelial permeability | 0.326 | 0.372 | 0.372 |
| E-selectin | SELE | Inflammation | 0.465 | 0.455 | 0.468 |
| G-CSF | CSF3 | Inflammation | 0.168 | 0.125 | 0.168 |
| ICAM-1 | ICAM1 | Inflammation | 0.285 | 0.291 | 0.273 |
| IL-10 | IL10 | Inflammation | 0.005 | -0.013 | 0.022 |
| IL-17A | IL17A | Inflammation | 0.044 | -0.001 | 0.001 |
| IL-18 | IL18 | Inflammation | 0.250 | 0.284 | 0.247 |
| IL-1B | IL1B | Inflammation | 0.189 | 0.221 | 0.219 |
| IL-6 | IL6 | Inflammation | 0.534 | 0.554 | 0.557 |
| IL-8 | CXCL8 | Inflammation | -0.047 | -0.044 | -0.049 |
| MPO | MPO | Inflammation | 0.374 | 0.410 | 0.391 |
| NAMPT | NAMPT | Inflammation | 0.309 | 0.361 | 0.371 |
| TNF | TNF | Inflammation | 0.072 | 0.066 | 0.044 |
| elafin | PI3 | Inflammation | 0.175 | 0.200 | 0.178 |
| elastase | ELANE | Inflammation | 0.270 | 0.334 | 0.324 |
| sTNFR-1 | TNFRSF1A | Inflammation | 0.326 | 0.405 | 0.405 |
| sTNFR-2 | TNFRSF1B | Inflammation | 0.286 | 0.317 | 0.307 |
| suPAR | PLAUR | Inflammation | 0.225 | 0.256 | 0.227 |
| BNP | NPPB | Myocardial | -0.103 | -0.110 | -0.146 |
| troponin I | TNNI3 | Myocardial | 0.067 | 0.019 | 0.004 |
| troponin T | TNNT2 | Myocardial | -0.029 | -0.015 | 0.015 |

*Values are the difference in standardized concentration between a body mass index of 35 and 25. Primary is the full cohort. The metabolically healthy stratum required no recorded occurrence of type 2 diabetes, hypertension or dyslipidemia, 5,327 participants, and is shown again restricted to the 3,881 with at least one diagnosis record.*

Supplementary dataset. Estimates for all 5,420 assays.

**E15. Reporting checklist**

The study is reported according to the STROBE recommendations for cross sectional studies. Table E9 gives the location of each item. Locations are named by section rather than by page, so they remain correct as the manuscript is typeset.

**Table E9. STROBE checklist for cross sectional studies.**

| **Section** | **Item** | **Recommendation** | **Location in this report** |
| --- | --- | --- | --- |
| Title and abstract | 1a | Study design indicated with a commonly used term in the title or abstract | Abstract, Methods |
|  | 1b | Informative and balanced summary of what was done and what was found | Abstract |
| Introduction | 2 | Background and rationale | Introduction, paragraphs 1 and 2 |
|  | 3 | Objectives, including prespecified hypotheses | Introduction, final paragraph |
| Methods | 4 | Key elements of study design | Methods, Data source |
|  | 5 | Setting, locations, and relevant dates of data collection | Methods, Data source |
|  | 6a | Eligibility criteria, sources and methods of selection of participants | Methods, Analytic cohort; Figure 1A |
|  | 7 | Outcomes, exposures, predictors, confounders and effect modifiers | Methods, Exposure and covariates |
|  | 8 | Sources of data and methods of measurement for each variable | Methods, Selected proteins and measurement; Methods, Exposure and covariates; Supplement E2 and E4 |
|  | 9 | Efforts to address potential sources of bias | Methods, Analytic cohort; Supplement E3 and E5 |
|  | 10 | How the study size was arrived at | Methods, Analytic cohort; Figure 1A |
|  | 11 | How quantitative variables were handled in the analyses | Methods, Statistical analysis; Supplement E8 |
|  | 12a | Statistical methods, including those used to control for confounding | Methods, Statistical analysis |
|  | 12b | Methods used to examine subgroups and interactions | Methods, Secondary analyses; Supplement E13 |
|  | 12c | How missing data were addressed | Methods, Exposure and covariates; Supplement E5 |
|  | 12d | Analytical methods taking account of the sampling strategy | Methods, Analytic cohort; Supplement E3 |
|  | 12e | Sensitivity analyses | Methods, Sensitivity analyses; Supplement E14; Table E7 |
| Results | 13a | Numbers of individuals at each stage of the study | Figure 1A; Results, first paragraph |
|  | 13b | Reasons for non-participation at each stage | Figure 1A |
|  | 13c | Consider use of a flow diagram | Figure 1A |
|  | 14a | Characteristics of participants and information on exposures and potential confounders | Table 1 |
|  | 14b | Number of participants with missing data for each variable of interest | Table 1 |
|  | 15 | Report numbers of outcome events or summary measures | Results; Figure 2; Table E4 |
|  | 16a | Unadjusted and confounder adjusted estimates and their precision | Results; Table 2; Table E4; Table E7 |
|  | 16b | Category boundaries when continuous variables were categorized | Methods, Statistical analysis; Figure 1B |
|  | 16c | Translation of relative risk into absolute risk for a meaningful time period | Not applicable, no risk estimates are reported |
|  | 17 | Other analyses done, such as subgroups, interactions and sensitivity analyses | Results; Tables E3, E5, E6, E7 and E8 |
| Discussion | 18 | Key results with reference to study objectives | Discussion, first paragraph |
|  | 19 | Limitations, including sources of potential bias or imprecision | Discussion, limitations paragraph |
|  | 20 | Cautious overall interpretation considering objectives, limitations, multiplicity and other evidence | Discussion |
|  | 21 | Generalisability of the study results | Discussion, limitations paragraph |
| Other information | 22 | Source of funding and the role of the funders | Funding |

Locations refer to the manuscript unless a supplement section or table is named. Item 16c is not applicable because the study reports differences in protein concentration across body size rather than risks of an event.
